# Somatic yoga therapy for functional neurological disorder: An experimental pilot study examining cognitive and affective mechanisms

**DOI:** 10.64898/2026.06.26.26356668

**Authors:** L. S. Merritt Millman, Emily Kennedy-Barnes, Anel Duarte, Jenna Pacelli, Yasmine Basamh, John Hodsoll, Susannah Pick

**Affiliations:** Institute of Psychiatry, Psychology & Neuroscience, King’s College London, London, UK

## Abstract

Accumulating evidence suggests alterations in neurocognitive, affective, interoceptive and autonomic processing in functional neurological disorder (FND), yet interventions targeting these processes remain underexplored. This study investigated the possible immediate and longer-term effects of a somatic yoga intervention on cognitive control, emotion regulation, state dissociation and affect, autonomic arousal, and interoceptive processing in FND. Twenty-three adults with FND completed six weeks of somatic yoga (N=12) or six weeks of a music-based relaxation control (N=11). At baseline, post-single session, and post-six weeks, participants completed laboratory measures of sustained attention, response inhibition, interoception, emotion regulation, and state dissociation and affect. Electrocardiography and galvanic skin conductance were recorded throughout. Linear mixed effects models assessed potential change on day one, immediately pre/post a single session, and from day 1 to the end of the six-week programme. After one session, stop signal reaction time, negative affect, and heartrate decreased in both groups (Δ=.69-.75). After one session and at six weeks, improved sustained attention, elevated positive affect, and reduced dissociation were seen in both groups, with a larger magnitude of change in yoga (Δ=.50-1.10). The yoga group exhibited fewer direction errors on the response inhibition task and shorter response times on the sustained attention task, with the opposite seen in the music group (Δ=.50-1.17). Both in the short- and longer-term, somatic yoga might lead to adaptive changes in attention and executive functioning, arousal, state affect and dissociation.

## Introduction

Functional neurological disorder (FND) is a condition situated at the interface of neurology and psychiatry, characterised by altered functioning of brain networks involved in movement, sensation, and cognition (APA, 2013). Individuals diagnosed with FND can experience motor symptoms, seizures, cognitive difficulties, or sensory alterations, with a marked impact on psychological and physical wellbeing, autonomy, and general functioning in everyday life (Hallett et al., 2022). The disorder may arise from a combination of psychological, biological, and social factors including comorbid mental health diagnoses, adverse life events, physical injuries and illnesses, or environmental stressors (Ludwig et al., 2018; Millman, Bhuma, et al., 2026; Millman et al., 2024; Pick et al., 2017).

Recent accounts of FND point towards alterations across attention, emotion regulation, and bodily awareness or interoceptive processing (Drane et al., 2021; Hallett et al., 2022; Pick et al., 2019). Reduced trust in the body and a heightened tendency to distract from unpleasant sensations (Millman et al., 2023; Pick et al., 2020; Stoffel, 2025), elevated psychological and somatoform dissociation (Brown & Reuber, 2016; Campbell et al., 2022; Pick et al., 2017), altered emotional regulation or responsivity (Pick et al., 2019; Pick et al., 2024), deficits in sustained attention and set-shifting (Pick et al., 2026), and autonomic dysregulation (Goldstein & Mellers, 2006; Paredes-Echeverri et al., 2022; Pick et al., 2019; Pick et al., 2017; Pick et al., 2024) have been found across FND subtypes. Further, disrupted integration of affective, cognitive, and sensory/interoceptive systems may play a role in the experience of functional neurological symptoms (Drane et al., 2021; Pick et al., 2019).

Therapeutics or interventions that work to enhance affective, functioning, autonomic regulation, and interoceptive processing in FND warrant exploration (Pick et al., 2019). Given the breathwork, accessible and gentle movement, and guided interoceptive or sensory attention involved in somatic yoga, this type of therapeutic could be of potential benefit in this population (Zaccaro et al., 2018). Somatic yoga may help to reduce sympathetic arousal (Field, 2016; Sullivan et al., 2018), encourage a mindful noticing of bodily sensations (Gibson, 2019; Lima-Araujo et al., 2022), enhance cognitive control (Gothe et al., 2019; Gothe & McAuley, 2015; Voss et al., 2023), and strengthen top-down regulation of affective and bodily states (Field, 2016; Gothe et al., 2019; Park et al., 2020; Streeter et al., 2012). Previous research has also documented the wellbeing and quality-of-life benefits of yoga, and reductions in other physical symptoms including pain and fatigue, which are often impacted in FND (Kipnis et al., 2024; Mooventhan & Nivethitha, 2017; Ross & Thomas, 2010).

Alongside primary feasibility measures and secondary physical and psychological outcomes reported in Kennedy-Barnes et al. (Kennedy-Barnes et al., 2026), this pilot study investigated the potential immediate (single session) and longer-term (six-week) effects of a somatic yoga intervention on state dissociation and affect, autonomic arousal, cognitive control, emotion regulation, interoceptive accuracy, and interoceptive confidence. In comparison to an active music-based relaxation control, the aim of this laboratory element was to gain insight regarding potential mechanisms involved in the somatic yoga intervention, to inform future larger-scale trials.

## Methods

### Study design and participants

This is a single-site, parallel-arm, prospectively registered (ISRCTN73085690) feasibility randomised controlled trial, designed to compare a six-week individual somatic yoga intervention with a six-week music-based relaxation programme. The King’s College London Health Faculties High Risk Research Ethics Committee (ref: HR/DP-24/25-46075), granted ethical approval, and the study was conducted in the Neurological Affective and Dissociative Symptoms (NEUROADS) Lab at the Institute of Psychiatry, Psychology & Neuroscience (IoPPN), King’s College London. Data collection occurred between March – June 2025.

Participant eligibility criteria are listed in Box 1. All participants self-referred to the study, either via online advertisements posted on King’s College London research volunteer pages, FND Hope UK, or relevant Facebook and Instagram groups, or after direct invitations sent to individuals previously consenting to be contacted regarding future studies in the NEUROADS Lab (secure registry held by the senior author, SP). Written, informed consent was provided by all participants prior to their enrolment in the study. Post-enrolment, participants completed a demographics and medical history screening interview, to confirm eligibility.

##### Box 1. Eligibility criteria.

| Inclusion criteria | Exclusion criteria |
| --- | --- |
| • 18 years or older<br>• Normal or corrected vision<br>• Fluency in English<br>• Formal diagnosis of FND made by a clinician (seizure, motor, or sensory symptoms) - established indirectly through documentation | • Major psychiatric, neurological, or cardiovascular disorders (e.g., active psychosis, severe substance use, epilepsy, heart disease)<br>• Purely functional cognitive symptoms<br>• Physical factors that may have limited or prevented participation (i.e., limb paralysis, more than 10 seizures/day, severe tremor)<br>• Current participation in/engagement with other body or music-based therapeutic interventions unable to be stopped across the study period |

A computer-generated allocation sequence was used to randomize participants 1:1 to either the somatic yoga intervention or music relaxation control (further detailed in (Kennedy-Barnes et al., 2026)). The first author (LSMM), who conducted all quantitative analyses, was blinded to group allocation throughout the study process.

### Interventions

Full details of both interventions are provided in Kennedy-Barnes et al. (Kennedy-Barnes et al., 2026), with manuals and practice logs included in the Supplementary Materials.

The somatic yoga intervention consisted of six, one-hour individual online sessions with the somatic yoga therapist (EKB). The intervention involved practices designed to enhance autonomic regulation and body awareness. Slow, gentle movements, breathing techniques (e.g., paced and diaphragmatic breathing), sensory-awareness exercises (e.g., eye-tracking), motor exploration (e.g., grounding, balance), and restorative postures (e.g., Savasana) were used within the intervention to encourage adaptive brain-body integration. A digital manual, practice log, and music playlist were provided for home practice, which was recommended 3-5 times/week. An audiovisual guide was incorporated following participant feedback in the second week of participation, to minimise cognitive demands and support accessibility when completing home practice.

Participants in the control arm engaged in 30–60-minute self-guided music listening, three-five times/week, across a six-week period. Participants were provided with three curated, instrumental playlists, one of which was identical to the playlist included in the yoga intervention. Slow, simple music, without much change in rhythm, was used across the three playlists to encourage activation of the parasympathetic nervous system and engage relevant neural circuits, without the inclusion of any guided bodily or breath-based practices. As with the yoga intervention, a digital manual and practice log, were provided for home practice. Check-ins once a week with EKB were equivalent to the yoga group regarding structure and amount of contact.

### Laboratory measures

Throughout the laboratory session, electrocardiography (ECG: electrodes on the left arm, right arm, and left leg in an Einthoven triangle) was used to measure heartrate and heartrate variability (RMSSD), and skin conductance was recorded during specific parts of the session using electrodermal activity sensors (EDA: Ag-AgCl electrodes with electrode paste, one on the distal phalange of the middle finger and the second on the distal phalange of the index finger of the non-dominant hand). These were recorded using a PowerLab data acquisition system and LabChart software (https://www.adinstruments.com/; see (Pick et al., 2016; Pick et al., 2018; Pick et al., 2024) for more detail).

Subjective momentary assessments of dissociation (Clinician Administered Dissociative States Scale, CADSS (Bremner et al., 1997)) and affect (positive and negative, Positive and Negative Affect Schedule, PANAS (Watson et al., 1988)) were measured four times during each laboratory session, in a counterbalanced order. These were administered using E-Prime experimental software (Psychology Software Tools, https://pstnet.com/products/e-prime/). The CADSS was used to measure present state dissociation (e.g., depersonalisation, derealisation, amnesia) across 19 self-reported items. The PANAS measured current state positive (10 items, e.g., interested, excited, proud) and negative (10 items, e.g., ashamed, hostile, afraid) affect across 20 self-report items.

Cognitive control was measured with two tasks from the Cambridge Neuropsychological Test Automated Battery (CANTAB) Connect (Cambridge Cognition, 2019), a touchscreen adaptation of the original CANTAB battery (Robbins et al., 1998; Robbins et al., 1994). The *Rapid Visual Information Processing Task* (RVIP) is a seven-minute test used to assess sustained attention. Participants are asked to detect target sequences of numbers (e.g., 3-5-7) within a much longer list of sequentially presented numbers (100 per minute) in a white box in the centre of the screen. If a participant has detected the target sequence, they are asked to press a button at the bottom of the screen as quickly as possible to indicate this. Key outcomes from this measure include response latency, ability score, probability of hit, total misses, and false alarm. The *Stop Signal Task* (SSRT) is a 14-minute test used to assess response inhibition. Participants are presented with an individual arrow, pointing either to the left or right of the screen. When the arrow appears, participants are asked to press the corresponding onscreen button (left or right) as quickly as possible. However, on some trials, participants will hear an auditory tone. In these cases, participants must withhold the button press. Key outcomes are errors (go trials), errors (stop trials), missed trials, and stop signal reaction time. These two tasks were administered in a counterbalanced order across participants and were selected for inclusion based on previous evidence of deficits in these domains in FND (Alluri et al., 2020; Millman et al., 2025; Pick et al., 2019; Pick et al., 2026; Van Patten et al., 2024).

The *Heartbeat Tracking Task* (HTT (Schandry, 1981)), used to measure interoceptive accuracy and confidence, was adapted from previous task versions used in the NEUROADS Lab (Millman et al., 2023; Pick et al., 2020) and administered using E-Prime. ECG was used to record actual heartbeats during task performance. The task involved counting the number of heartbeats felt over varying amounts of time (undisclosed to participants). Standardised written instructions were provided onscreen (detailed in Table S1; (Millman, Davies, et al., 2026)). Two task versions (25, 30, 35, and 40s or 29, 34, 39, and 44s) were designed, with the order counterbalanced across participants. Participants were asked to only count heartbeats that they felt, with clear instructions not to count seconds, take a guess or check their pulse during the task (Desmedt et al., 2020; Millman et al., 2023; Pick et al., 2020). Participants manually entered the number of heartbeats they had felt after completion of each trial (indicative of interoceptive accuracy). They were also asked to rate how confident they were with regards to the number of heartbeats felt during each trial on a scale from 0-10 (low-high certainty; indicative of state interoceptive sensibility). Rest periods in between each trial were included, after reporting the above, as well as one practice trial at the beginning of the task. Trial durations were not disclosed, and participants were not provided with any feedback on how they performed.

An affective images task, administered using E-Prime experimental software and adapted from previous task versions used in the NEUROADS Lab (Pick et al., 2025; Pick et al., 2024) was included as a measure of emotional response and regulation. Affective images (negative and neutral) were selected(Lang et al., 2005) based on normative arousal and valence ratings. Image blocks across types were roughly matched for content, and blocks within the same image type were matched for normative valence and arousal rating. Two versions of the task were developed, with the same images used in both versions, but presented in varying blocks. For each image block, participants were instructed to either simply observe their responses to the presented images (Watch) or try to down-regulate any emotional responses to the images (Dampen), with dampening strategies reported(Pick et al., 2024).

Four blocks of each of the following conditions were administered: Negative-Watch, Negative-Distance, Neutral-Watch. Participants viewed a total of 12 image blocks, with four images per block, and block order was pseudorandomised so that no more than two blocks of the same condition were successively presented. Standardised written instructions (Table S2) were presented onscreen throughout the task. Each block started with the relevant task instruction (Watch/Dampen) presented onscreen, followed by a central fixation cross, and then four images of the same type (Negative/Neutral) presented in a random order. As used in Pick, Mellers and Goldstein (Pick et al., 2018) and Pick et al. (Pick et al., 2024), immediately after each image block, participants were asked to complete momentary subjective assessments on digitised Self-Assessment Manikins (Lang et al., 2005) of valence (1 “unpleasant/unhappy” - 9 “pleasant/happy”) and arousal (1 “relaxed/calm” - 9 “tense/excited”), immediately followed by a rest period. Heartrate, heartrate variability, and skin conductance were continuously recorded throughout task completion.

### Procedure

After arriving for their first in-person visit at the IoPPN, participants began by completing the CADSS and PANAS tasks. Following this, ECG electrodes and EDA sensors were attached, and, after a 3-minute habituation period, a 5-minute recording of resting state psychophysiology was completed. EDA sensors were then removed to not interfere with cognitive task performance (use of both index fingers required for SSRT), and the CANTAB RVIP and SSRT were completed. These tasks were followed by administration of one version of the HTT. The EDA sensors were then replaced, and one version of the affective images task was run. At the end of this session, the CADSS and PANAS were completed for a second time. These procedures took approximately one hour to complete. After completion of this laboratory session, participants moved to another room to complete a single session of their assigned intervention (yoga or music) with the somatic yoga therapist (EKB). Upon completion of the intervention session, participants returned to the laboratory to repeat the same procedures described above. All laboratory sessions were led by SP or LSMM. Participants were reimbursed with a £30 shopping voucher at the end of their visit. After completing the six-week yoga or music programme, with online sessions once per week, participants returned for a second in-person visit. This involved the same initial laboratory session, followed by a final consolidation yoga or music session with EKB.

### Analysis

All analyses were conducted by LSMM using R and RStudio (Version 4.5.1), with statistical oversight from JH. Demographics are presented separately for each group in Table 1. Raw descriptive statistics for all laboratory measures are presented in Table S3, separated by arm and timepoint.

**Table 1.** Demographic and clinical characteristics across both groups.

|  | **Yoga**<br>**N=12**<br>**n (%)** | **Music**<br>**N=11**<br>**n (%)** |
| --- | --- | --- |
| **Gender (% female)** | 11 (92%) | 11 (100%) |
| **Sex (% female)** | 11 (92%) | 11 (100%) |
| **Age (M [SD])** | 45.9 (13.0) | 44.9 (11.6) |
| **Handedness (% right)** | 12 (100%) | 10 (91%) |
| **Smoking status (% yes)** | 3 (25%) | 0 (0%) |
| **Body Mass Index (M [SD])** | 29.7 (7.4) | 26.0 (5.2) |
| **Employment (% employed)** | 6 (50%) | 4 (36%) |
| **Education (% undergraduate or postgraduate)** | 6 (50%) | 7 (64%) |
| **Ethnicity** |  |  |
| *White* | 9 (75%) | 7 (64%) |
| *Black, Asian, or Mixed* | 3 (25%) | 4 (36%) |
| **Previous knowledge of heartrate** | 6 (50%) | 4 (36%) |
| **Current medication (% yes)** | 9 (75%) | 10 (91%) |
| **Current mental health diagnosis (% yes)** | 10 (83%) | 7 (64%) |
| **Current physical health diagnoses (% yes)** | 8 (67%) | 9 (82%) |
| **Clinical Global Impression – Severity (M, SD)** | 4.1 (0.9) | 3.6 (1.0) |
| **Primary self-reported FND symptoms** |  |  |
| *Mobility/gait difficulties* | 3 (25%) | 5 (45%) |
| *Sensory symptoms* | 3 (25%) | 2 (18%) |
| *Seizures* | 4 (33%) | 5 (45%) |
| *Tremors* | 3 (25%) | 2 (18%) |
| *Jerks/spasms/tics* | 5 (42%) | 3 (27%) |
| *Weakness/paralysis* | 4 (33%) | 4 (36%) |
| *Speech difficulties* | 2 (17%) | 2 (18%) |

Notes. M=Mean, SD=Standard deviation. \*Measured at baseline

Psychophysiological data (ECG, EDA) preprocessing involved visual screening of ECG and EDA data for artefacts, and sections of contaminated and/or poor-quality data were excluded before analysis. Using LabChart, average heartrate, heartrate variability (RMSSD), and skin conductance levels were extracted for the 5-minute baseline period recording, as well as for each block within the emotion regulation task (12 individual blocks). To calculate interoceptive accuracy, the proportional discrepancy between the actual and perceived number of heartbeats was calculated using the following formula: 1/4 ∑ [(1 − (|actual heartbeats – perceived heartbeats|/actual heartbeats)]. This resulted in an accuracy error index from 0-1 with values closer to 1 reflecting a lower discrepancy and superior interoceptive accuracy.

Linear mixed effects models were run to calculate effect sizes (contrast from each mixed model / baseline SD of music control group; Glass’s Δ) using the lme4 and emmeans packages (Lenth, 2023). Due to the aims of this pilot study, separate models were set up to assess: 1) potential change due to the yoga intervention on day one, immediately pre/post a single session of yoga, and 2) assessing potential change due to the yoga intervention from day 1 to the end of the programme at visit 2.

For measures of cognitive control, autonomic arousal, and interoception, Group (yoga intervention, music control), Time (baseline, post-single session of yoga/music OR baseline, post-six week yoga/music programme), and an interaction term between Group and Time were included as fixed effects, and Participant as random effect on the following variables: HTT accuracy, HTT confidence, heartrate, skin conductance, heartrate variability, response latency, ability score, probability of hit, total misses and false alarm on the RVIP, errors (go trials), errors (stop trials), missed trials, and stop signal reaction time on the SST (Table S4, S8). Models were run separately to assess any main effects or interactions pre-post a single session of yoga or music, as well as pre-post the six-week programme. The music group and baseline scores were the reference levels within these analyses.

For state dissociation and affect, Group (yoga intervention, music control), Time (visit 1 pre-single yoga/music session, visit 1 post-single yoga/music session OR visit 1 pre-single yoga/music session, post six-week yoga/music programme), and an interaction term between Group and Time were included as fixed effects, and Participant as random effect on the following variables: positive affect, negative affect, state dissociative symptoms (Table S4, S8). Models were run separately to assess any main effects or interactions immediately pre-post a single session of yoga or music, as well as pre-post the six-week programme. The music group and visit 1 pre single yoga/music session were the reference levels within these analyses.

For the emotion regulation task, Group (yoga intervention, music control), Time (baseline, post-single session of yoga/music OR baseline, post-six week yoga/music programme), Condition (Neutral, Negative-Watch, Negative-Dampen), and an interaction term between Group, Time and Condition were included as fixed effects, and Participant as random effect on the following variables: valence, subjective arousal, heartrate, heartrate variability (RMSSD), and electrodermal activity (Table S4, S8). The music group and baseline scores were the reference levels within these analyses.

Rates of missing data were calculated and presented for each variable across all timepoints (Table S5). Generally, linear mixed models can handle unbalanced designs including missing data due to the estimation of model parameters using observed data (Hox et al., 2017).

Exploratory calculations examined interoceptive insight (the correlation between HTT accuracy and HTT confidence) at each time point (baseline, post single session, end intervention), separately within each group (yoga, music), using Pearson’s (normal) or Spearman’s (correlations), as required. Correlations between dissociation (CADSS), affect (positive, negative), autonomic states (heartrate, RMSSD, skin conductance), and heartbeat tracking accuracy and confidence were also calculated and explored within groups at each timepoint.

## Results

### Demographics

The participant groups were matched in age, sex, gender, and handedness (Table 1), and were matched regarding current self-reported medication, mental health diagnoses, and physical health diagnoses. Average body mass index was slightly elevated in the yoga group. The types of primary FND symptoms reported across both groups included mobility/gait difficulties, sensory symptoms, seizures, tremors, jerks, weakness/paralysis, and speech difficulties.

### Laboratory measures

Consistent with Kennedy-Barnes et al. (Kennedy-Barnes et al., 2026), these results focus on effect sizes (medium-large, Δ≥.50) as an indicator of potential signal to inform future, larger-scale trials.

### Pre-post single session of yoga or music

No medium-large effects for Group x Time x Condition interactions were found across all variables (Table S6).

#### Cognitive control

After a single session of yoga or music, regardless of group, stop signal reaction time (SSTSSRT; Δ=.69) decreased (Table 2, S7). Regardless of timepoint, the music group reported shorter stop signal reaction times (SSTSSRT; Δ=1.39, 95% CI: -2.71, 42.41), and fewer direction errors on stop trials (SSTDES; Δ=1.51) relative to the yoga group.

**Table 2.**
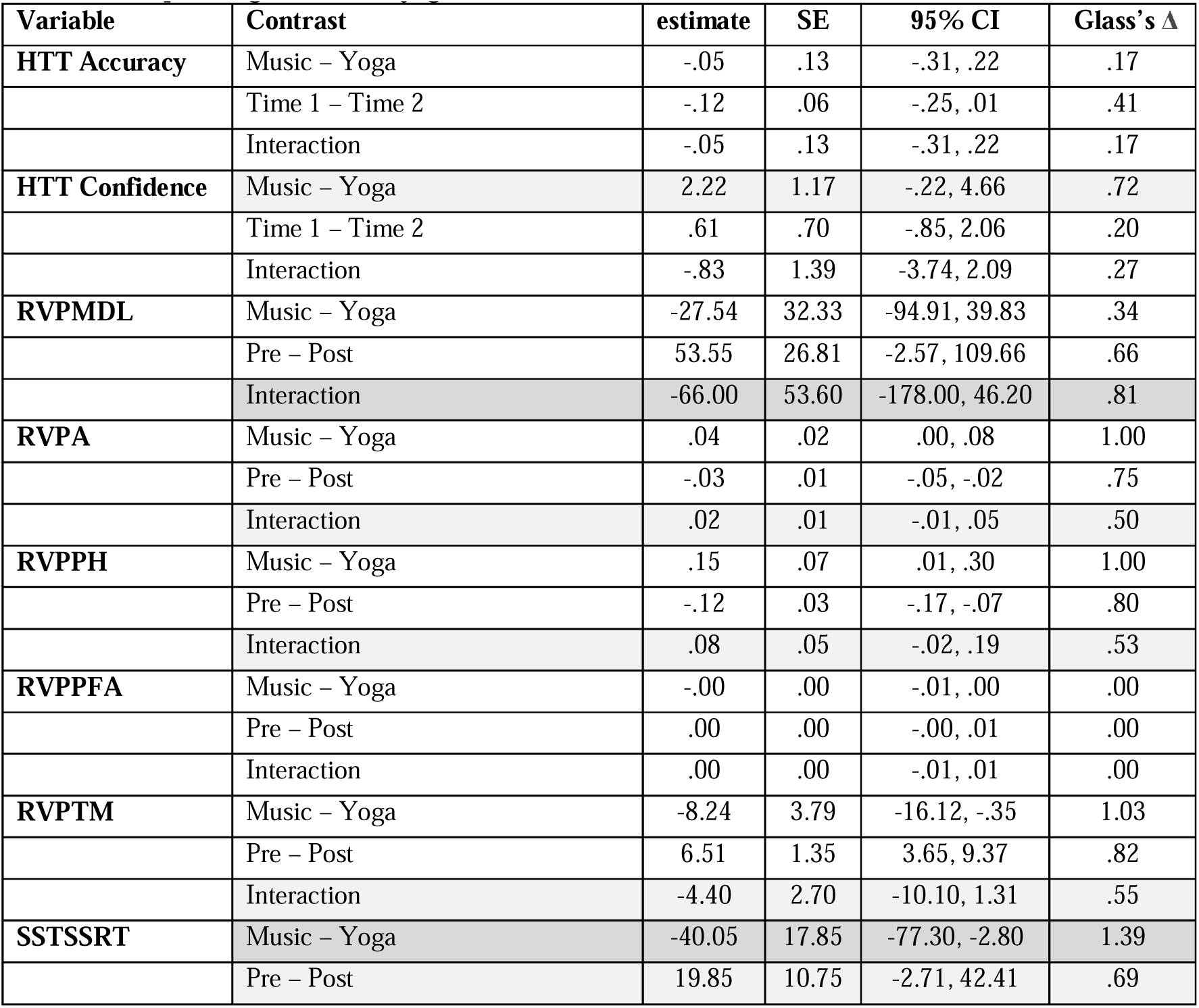
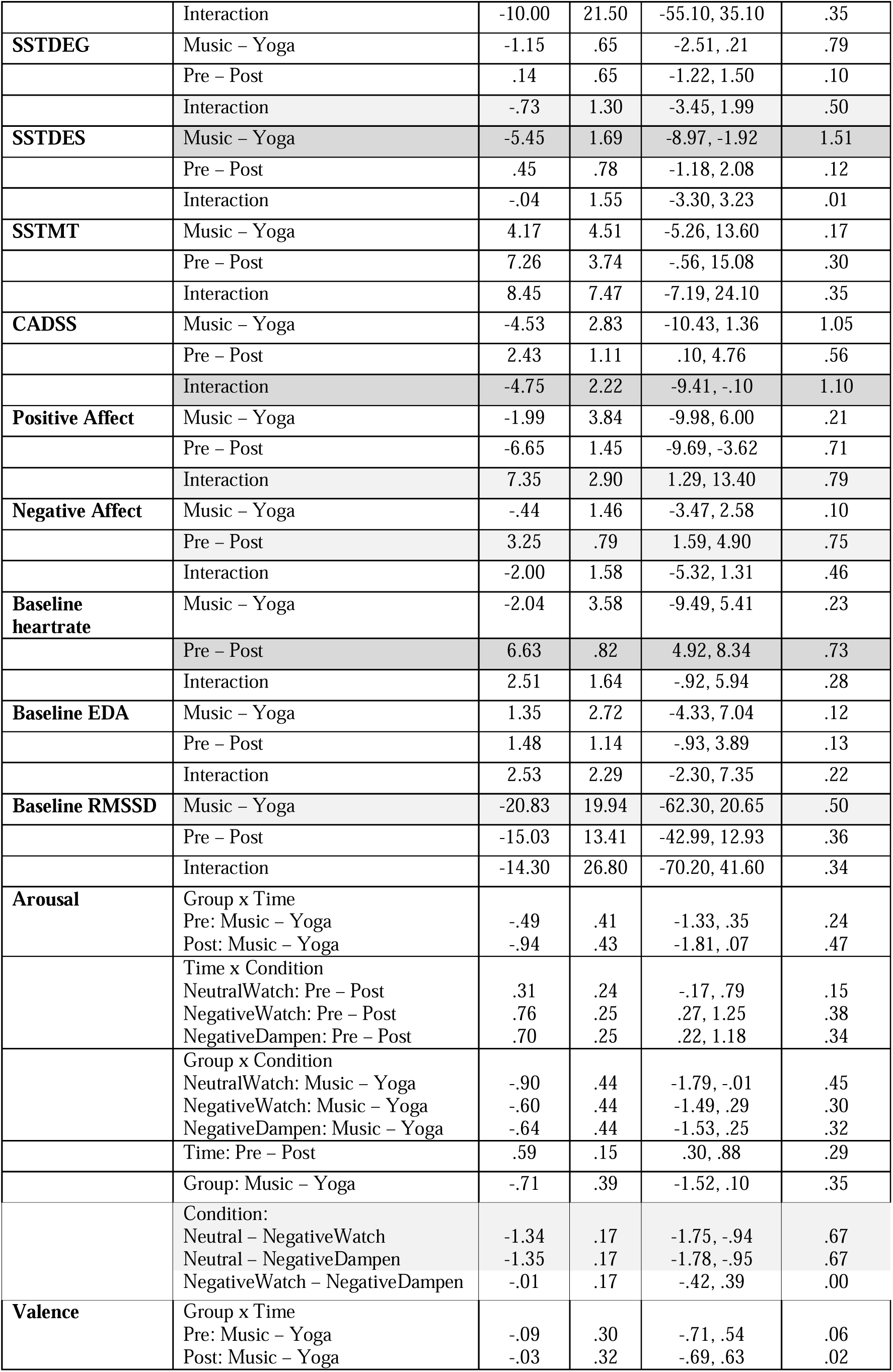
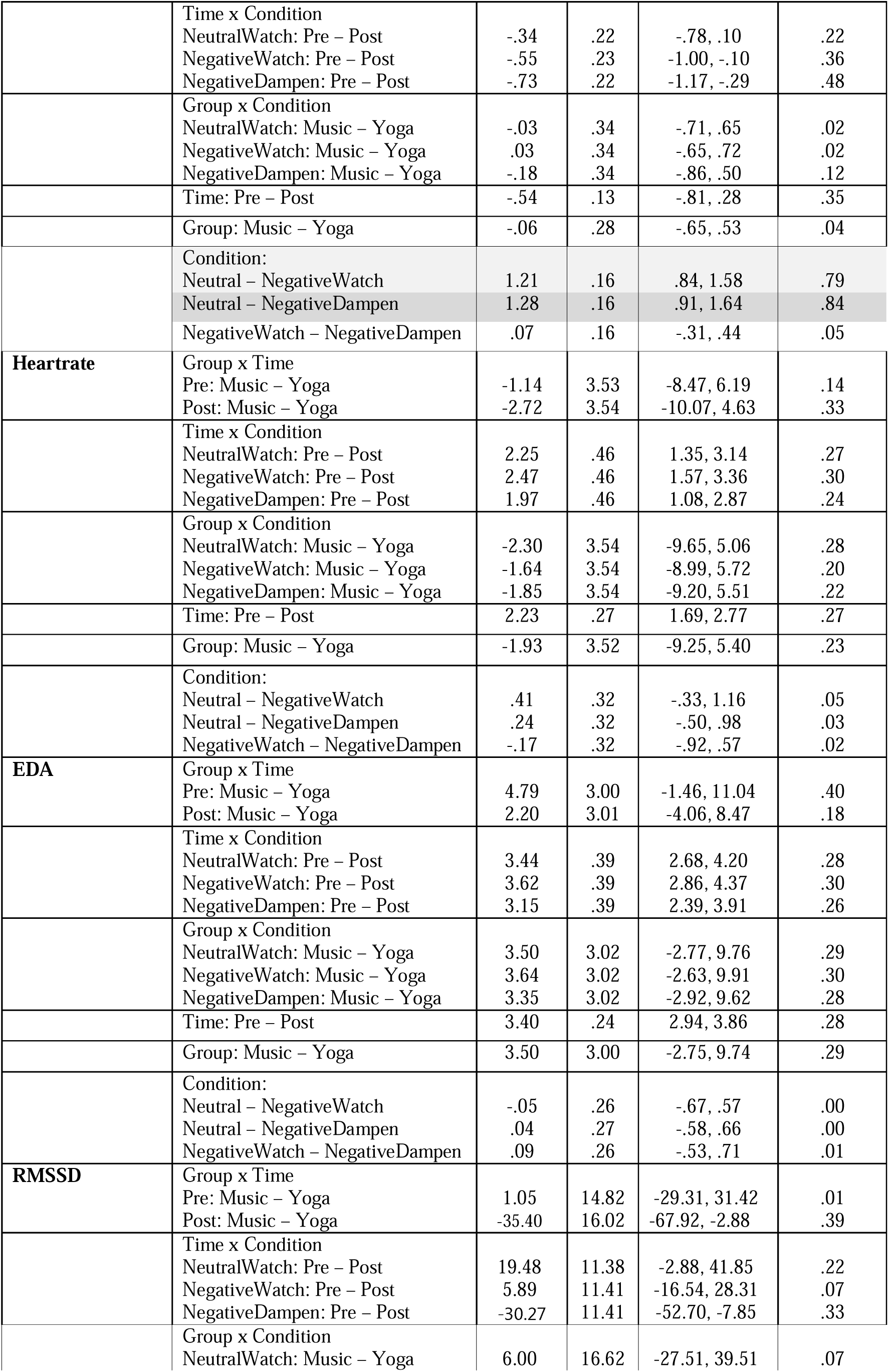
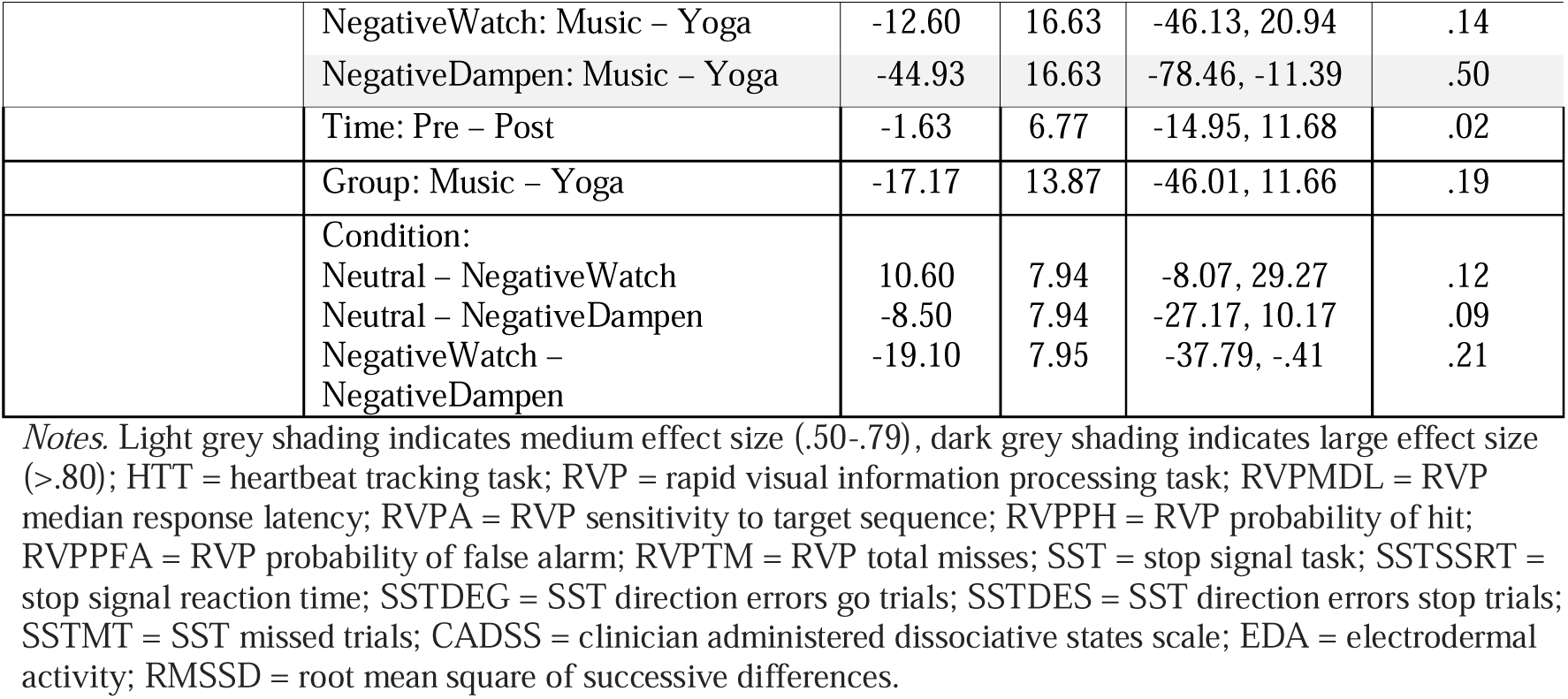
Pre-post single session of yoga or music. (Linear Mixed Model emmeans contrasts).

Medium-large effect sizes for Group x Time interactions, suggestive of differing magnitudes of change between the two groups, were present for RVPA (Δ=.50; 95% CI: -.01, .05), RVPPH (Δ=.53; 95% CI: -.02, .19), RVPMDL (Δ=.81; 95% CI: -178.00, 46.20), and RVPTM (Δ=.55; 95% CI: -10.10, 1.31). Both groups exhibited increases (RVPA, RVPPH) or reductions (RVPMDL, RVPTM) on these measures from pre-post single session of yoga or music, but larger changes were seen in the yoga group.

For direction errors (go trials; SSTDEG) on the SST, estimated marginal means suggested divergent trajectories across time between the two groups, with fewer errors in the yoga group, and more in the music group, after a single session of the intervention (Δ=.50, 95% CI: -3.45, 1.99; Table 2, S7). It is important to note that the standard error (SE=1.30) is large relative to the estimate of group difference (-.73) for this measure.

#### Interoceptive accuracy and confidence

Regardless of timepoint, the music group reported higher levels of confidence in the HTT (Δ=.72, 95% CI: -.22, 4.66).

#### State dissociation and affect

After a single session of yoga or music, regardless of group, negative affect (Δ=.75) decreased (Table 2, S7). Medium-large effect sizes for Group x Time interactions, suggestive of differing magnitudes of change between the two groups, were present for CADSS (Δ=1.10) and positive affect (Δ=.79; Table 2, S7). Both groups exhibited increases (positive affect) or reductions (CADSS) on these measures from pre-post single session of yoga or music, but larger changes were seen in the yoga group.

#### Emotion regulation and autonomic arousal

Regardless of timepoint and group, subjective arousal scores were elevated in the Negative-Watch and Negative-Dampen conditions of the affective images task (Δ=.67), and valence was rated as less pleasant in the same conditions (Δ=.79-.84), relative to the Neutral condition (Table 2, S7). In the Negative-Dampen condition and at baseline, regardless of timepoint, the music group exhibited lower RMSSD (Δ=.50) relative to the yoga group. Regardless of group, average baseline heartrate was reduced after a single session of yoga or music (Δ=.73).

### Pre-post six weeks of yoga or music

No medium-large effects for Group x Time x Condition interactions were found across all variables (Table S9).

#### Cognitive control

Regardless of timepoint, the music group exhibited faster reaction times (SSTSSRT, Δ=1.43) on the SST, relative to the yoga group (Table 3, S10).

**Table 3.** Pre-post six weeks of yoga or music. (Linear Mixed Model emmeans contrasts).

| Variable | Contrast | estimate | SE | 95% CI | Glass's $\Delta$ |
| --- | --- | --- | --- | --- | --- |
| **HTT Accuracy** | Music – Yoga | -.10 | .12 | -.36, .16 | .34 |
|  | Pre – Post | -.13 | .07 | -.27, .01 | .45 |
|  | Interaction | .06 | .13 | -.23, .34 | .21 |
| **HTT Confidence** | Music – Yoga | 1.28 | 1.28 | -1.39, 3.95 | .41 |
|  | Pre – Post | -.06 | .48 | -1.07, .95 | .02 |
|  | Interaction | 1.06 | .96 | -.97, 3.08 | .34 |
| **RVPMDL** | Music – Yoga | -13.08 | 37.34 | -91.05, 64.88 | .16 |
|  | Pre – Post | 33.29 | 33.86 | -37.67, 104.24 | .41 |
|  | Interaction | -95.20 | 67.80 | -237.00, 46.70 | 1.17 |
| **RVPA** | Music – Yoga | .03 | .02 | -.01, .07 | .75 |
|  | Pre – Post | -.04 | .01 | -.05, -.02 | 1.00 |
|  | Interaction | .04 | .01 | .01, .07 | 1.00 |
| **RVPPH** | Music – Yoga | .10 | .07 | -.05, .26 | .67 |
|  | Pre – Post | -.14 | .02 | -.20, -.09 | .93 |
|  | Interaction | .17 | .05 | .07, .28 | 1.13 |
| **RVPPFA** | Music – Yoga | -.00 | .00 | -.01, .00 | .00 |
|  | Pre – Post | .00 | .00 | -.00, .00 | .00 |
|  | Interaction | .00 | .00 | -.00, .01 | .00 |
| **RVPTM** | Music – Yoga | -5.62 | 3.96 | -13.86, 2.62 | .70 |
|  | Pre – Post | 7.74 | 1.32 | 4.94, 10.54 | .97 |
|  | Interaction | -9.36 | 2.64 | -15.00, -3.76 | 1.17 |
| **SSTSSRT** | Music – Yoga | -41.40 | 17.98 | -78.81, -3.98 | 1.43 |
|  | Pre – Post | 11.68 | 10.89 | -11.17, 34.52 | .40 |
|  | Interaction | -6.89 | 21.80 | -52.60, 38.80 | .24 |
| **SSTDEG** | Music – Yoga | -.63 | .72 | -2.12, .87 | .43 |
|  | Pre – Post | .51 | .63 | -.80, 1.83 | .35 |
|  | Interaction | -1.82 | 1.26 | -4.45, .81 | 1.26 |
| **SSTDES** | Music – Yoga | -4.43 | 1.15 | -6.82, -2.03 | 1.22 |
|  | Pre – Post | -1.06 | .79 | -2.70, .59 | .29 |
|  | Interaction | -2.11 | 1.57 | -5.39, 1.18 | .58 |
| **SSTMT** | Music – Yoga | 8.97 | 6.45 | -4.43, 22.38 | .37 |
|  | Pre – Post | 3.38 | 1.79 | -.41, 7.16 | .14 |
|  | Interaction | -3.12 | 3.58 | -10.70, 4.45 | .13 |
| **CADSS** | Music – Yoga | -5.82 | 2.44 | -10.90, -.75 | 1.35 |
|  | Pre – Post | 3.14 | 1.55 | -.12, 6.40 | .73 |
|  | Interaction | -2.17 | 3.10 | -8.69, 4.35 | .50 |
| **Positive Affect** | Music – Yoga | -1.68 | 3.37 | -8.68, 5.32 | .18 |
|  | Pre – Post | -6.26 | 1.75 | -9.95, -2.57 | .67 |
|  | Interaction | 6.74 | 3.51 | -.64, 14.10 | .72 |
| **Negative Affect** | Music – Yoga | -1.68 | 1.62 | -5.06, 1.70 | .39 |
|  | Pre – Post | .94 | 1.41 | -2.01, 3.89 | .22 |
|  | Interaction | .47 | 2.82 | -5.43, 6.36 | .11 |
| **Baseline Heartrate** | Music – Yoga | .44 | 4.21 | -8.32, 9.20 | .05 |
|  | Pre – Post | 1.01 | 2.05 | -3.30, 5.32 | .11 |
|  | Interaction | -2.45 | 4.11 | -11.10, 6.18 | .27 |
| **Baseline EDA** | Music – Yoga | 2.46 | 2.47 | -2.70, 7.63 | .22 |
|  | Pre – Post | 2.88 | 1.44 | -.15, 5.91 | .25 |
|  | Interaction | .87 | 2.88 | -5.18, 6.93 | .08 |
| **Baseline RMSSD** | Music – Yoga | -30.63 | 22.20 | -76.80, 15.54 | .73 |
|  | Pre – Post | -8.43 | 13.55 | -36.84, 19.97 | .20 |
|  | Interaction | 5.33 | 27.10 | -51.50, 62.10 | .13 |
| **Arousal** | Group x Time |  |  |  |  |
|  | Pre: Music – Yoga | -.50 | .36 | -1.23, .23 | .24 |
|  | Post: Music – Yoga | .01 | .38 | -.76, .78 | .00 |
|  | Time x Condition |  |  |  |  |
|  | NeutralWatch: Pre – Post | .11 | .27 | -.41, .64 | .05 |
|  | NegativeWatch: Pre – Post | .36 | .28 | -.18, .91 | .17 |
|  | NegativeDampen: Pre – Post | .60 | .27 | .07, 1.14 | .29 |
|  | Group x Condition |  |  |  |  |
|  | NeutralWatch: Music – Yoga | -.37 | .39 | -1.16, .43 | .18 |
|  | NegativeWatch: Music – Yoga | -.31 | .40 | -1.12, .50 | .15 |
|  | NegativeDampen: Music – Yoga | -.06 | .40 | -.86, .74 | .03 |
|  | Time: Pre – Post | .36 | .16 | .04, .68 | .17 |
|  | Group: Music – Yoga | -.24 | .33 | -.93, .45 | .12 |
|  | Condition: |  |  |  |  |
|  | Neutral – NegativeWatch | -1.45 | .19 | -1.90, -1.00 | .72 |
|  | Neutral – NegativeDampen | .16 | .19 | -1.73, -.85 | .08 |
|  | NegativeWatch – NegativeDampen | .16 | .19 | -.29, .61 | .08 |
| **Valence** | Group x Time |  |  |  |  |
|  | Pre: Music – Yoga | -.07 | .26 | -.60, .46 | .04 |
|  | Post: Music – Yoga | .21 | .28 | -.36, .79 | .13 |
|  | Time x Condition |  |  |  |  |
|  | NeutralWatch: Pre – Post | -.44 | .23 | -.89, -.00 | .27 |
|  | NegativeWatch: Pre – Post | -.17 | .23 | -.62, .28 | .10 |
|  | NegativeDampen: Pre – Post | -.62 | .22 | -1.06, -.18 | .37 |
|  | Group x Condition |  |  |  |  |
|  | NeutralWatch: Music – Yoga | .00 | .30 | -.60, .60 | .00 |
|  | NegativeWatch: Music – Yoga | .35 | .30 | -.26, .96 | .21 |
|  | NegativeDampen: Music – Yoga | -.14 | .30 | -.74, .46 | .08 |
|  | Time: Pre – Post | -.41 | .13 | -.67, -.15 | .25 |
|  | Group: Music – Yoga | .07 | .24 | -.42, .57 | .04 |
|  | Condition: |  |  |  |  |
|  | Neutral – NegativeWatch | 1.47 | .16 | 1.09, 1.84 | .96 |
|  | Neutral – NegativeDampen | 1.38 | .16 | 1.01, 1.75 | .90 |
|  | NegativeWatch – NegativeDampen | -.08 | .16 | -.46, .29 | .05 |
| **Heartrate** | Group x Time |  |  |  |  |
|  | Pre: Music – Yoga | -1.09 | 3.67 | -8.71, 6.53 | .13 |
|  | Post: Music – Yoga | .54 | 3.69 | -7.11, 8.19 | .07 |
|  | Time x Condition |  |  |  |  |
|  | NeutralWatch: Pre – Post | .66 | .65 | -.61, 1.94 | .08 |
|  | NegativeWatch: Pre – Post | .95 | .65 | -.32, 2.23 | .11 |
|  | NegativeDampen: Pre – Post | 1.40 | .64 | .14, 2.67 | .17 |
|  | Group x Condition |  |  |  |  |
|  | NeutralWatch: Music – Yoga | -.60 | 3.69 | -8.26, 7.07 | .07 |
|  | NegativeWatch: Music – Yoga | -.38 | 3.69 | -8.04, 7.28 | .05 |
|  | NegativeDampen: Music – Yoga | .16 | 3.69 | -7.50, 7.82 | .02 |
|  | Time: Pre – Post | 1.01 | .39 | .25, 1.77 | .12 |
|  | Group: Music – Yoga | -.27 | 3.66 | -7.88, 7.33 | .03 |
|  | Condition: |  |  |  |  |
|  | Neutral – NegativeWatch | .49 | .45 | -.57, 1.55 | .06 |
|  | Neutral – NegativeDampen | .78 | .45 | -.27, 1.84 | .09 |
|  | NegativeWatch – NegativeDampen | .30 | .45 | -.76, 1.35 | .04 |
| **EDA** | Group x Time |  |  |  |  |
|  | Pre: Music – Yoga | 4.37 | 2.92 | -1.70, 10.44 | .36 |
|  | Post: Music – Yoga | 1.83 | 2.93 | -4.24, 7.91 | .15 |
|  | Time x Condition |  |  |  |  |
|  | NeutralWatch: Pre – Post | 3.74 | .46 | 2.83, 4.66 | .31 |
|  | NegativeWatch: Pre – Post | 3.98 | .46 | 3.07, 4.89 | .33 |
|  | NegativeDampen: Pre – Post | 3.74 | .46 | 2.84, 4.65 | .31 |
|  | Group x Condition |  |  |  |  |
|  | NeutralWatch: Music – Yoga | 3.13 | 2.93 | -2.96, 9.22 | .26 |
|  | NegativeWatch: Music – Yoga | 3.17 | 2.93 | -2.91, 9.26 | .26 |
|  | NegativeDampen: Music – Yoga | 3.00 | 2.93 | -3.09, 9.09 | .25 |
|  | Time: Pre – Post | 3.82 | .28 | 3.27, 4.37 | .32 |
|  | Group: Music – Yoga | 3.10 | 2.91 | -2.95, 9.15 | .26 |
|  | Condition: |  |  |  |  |
|  | Neutral – NegativeWatch | -.01 | .32 | -.77, .74 | .00 |
|  | Neutral – NegativeDampen | .24 | .32 | -.51, .99 | .02 |
|  | NegativeWatch – NegativeDampen | .26 | .32 | -.49, 1.01 | .02 |
| **RMSSD** | Group x Time |  |  |  |  |
|  | Pre: Music – Yoga | .87 | 14.68 | -29.21, 30.95 | .01 |
|  | Post: Music – Yoga | -31.12 | 15.85 | -63.28, 1.03 | .34 |
|  | Time x Condition |  |  |  |  |
|  | NeutralWatch: Pre – Post | 23.36 | 11.29 | 1.18, 45.55 | .26 |
|  | NegativeWatch: Pre – Post | -.08 | 11.30 | -22.28, 22.13 | .00 |
|  | NegativeDampen: Pre – Post | -9.97 | 11.20 | -31.98, 12.04 | .11 |
|  | Group x Condition |  |  |  |  |
|  | NeutralWatch: Music – Yoga | 2.14 | 16.49 | -31.10, 35.39 | .02 |
|  | NegativeWatch: Music – Yoga | -18.37 | 16.50 | -51.64, 14.90 | .20 |
|  | NegativeDampen: Music – Yoga | -29.16 | 16.43 | -62.30, 3.98 | .32 |
|  | Time: Pre – Post | 4.44 | 6.66 | -8.65, 17.53 | .05 |
|  | Group: Music – Yoga | -15.13 | 13.75 | -43.70, 13.45 | .17 |
|  | Condition: |  |  |  |  |
|  | Neutral – NegativeWatch | 5.54 | 7.89 | -13.01, 24.08 | .06 |
|  | Neutral – NegativeDampen | -.43 | 7.85 | -18.89, 18.04 | .00 |
|  | NegativeWatch – NegativeDampen | -5.97 | 7.85 | -24.44, 12.50 | .07 |

*Notes.* Light grey shading indicates medium effect size (.50-.79), dark grey shading indicates large effect size (>.80); HTT = heartbeat tracking task; RVP = rapid visual information processing task; RVPMDL = RVP median response latency; RVPa = RVP sensitivity to target sequence; RVPPh = RVP probability of hit; RVPpfa = RVP probability of false alarm; RVPTM = RVP total misses; SST = stop signal task; SSTSSRT = stop signal reaction time; SSTDEG = SST direction errors go trials; SSTDES = SST direction errors stop trials; SSTMT = SST missed trials; CADSS = clinician administered dissociative states scale.

Medium-large effect sizes for Group x Time interactions, suggestive of differing magnitudes of change between the two groups, were present for RVPA (Δ=1.00), RVPPH (Δ=1.13), and RVPTM (Δ=1.17). Both groups exhibited increases (RVPA, RVPPH) or reductions (RVPTM) on these measures from pre-post six weeks of yoga or music, but larger changes were seen in the yoga group.

For median response latency on the RVP task (RVPMDL, Δ=1.17, 95% CI: -237.00, 46.70) and direction errors (go trials; SSTDEG, Δ=1.26, 95% CI: -4.45, .81, stop trials; SSTDES, Δ=.58; 95% CI: -5.39, 1.18) on the SST, estimated marginal means showed shorter response times and fewer errors in the yoga group, and the opposite in the music group, after six weeks of the intervention (Table 3, S10).

#### State dissociation and affect

Medium-large effect sizes for Group x Time interactions, suggestive of differing magnitudes of change between the two groups, were present for CADSS (Δ=.50; 95% CI: -8.69, 4.35), and positive affect (Δ=.72; 95% CI: -.64, 14.10; Table 3, S10). Both groups exhibited increases (positive affect) or reductions (CADSS) on these measures from pre-post six weeks of yoga or music, but larger changes were seen in the yoga group.

#### Emotion regulation and autonomic arousal

Regardless of timepoint and group, subjective arousal scores were elevated in the Negative-Watch condition of the affective images task (Δ=.72), and subjective valence was rated as less pleasant in the Negative-Watch and Negative-Dampen conditions (Δ=.90-.96), relative to the Neutral condition (Table 3, S10). At baseline, regardless of timepoint, the music group exhibited lower RMSSD (Δ=.73) compared to the yoga group.

## Discussion

This pilot study investigated possible immediate (single session) and longer-term (six-week) effects of an individual somatic yoga intervention compared to a music-based relaxation condition on cognitive control, state dissociation and affect, emotion regulation, autonomic arousal, interoceptive accuracy and interoceptive confidence in adults with FND. Preliminary results point towards changes in attention and executive functioning, autonomic arousal, and state dissociation and affect immediately, and/or in the longer-term, after completion of the somatic yoga intervention.

After a single session and maintained at six weeks, improvements in sustained attention on a validated CANTAB test were observed in both groups, including an elevated sensitivity to detect a target digit sequence, increased probability of detecting/responding to the target sequence, and reduced total misses of target sequences, with larger changes specific to the yoga group (Δ=.50-1.17). After a single session only, median response latency on the sustained attention test also improved in both groups, but at six weeks, this measure remained improved in yoga while worsening in the music group. These improvements in attention seen in the yoga group corroborate previous findings of enhanced attention and processing speed in RCTs of longer-term yoga interventions in healthy and clinical groups, and single session, acute exposures, in healthy groups (Gothe & McAuley, 2015). Schmalzl, Powers & Blom (Schmalzl et al., 2015) similarly summarised evidence for improved visual attention after a 10-day yoga program (Telles et al., 1995) and in yoga practitioners (Narayana, 2009).

Improved executive functioning has also been reported across previous studies (Gothe & McAuley, 2015), and in a systematic review of yoga-based interventions on cognitive function in older adults (working memory, mental shifting/flexibility; (Hoy et al., 2021)). Our study adds to this evidence base, with reductions in stop signal reaction time in both the yoga and music groups after a single session, and fewer direction errors specifically seen in the yoga group after a single session and maintained at six weeks (Δ=.50-1.26), diverging from the music group where more errors were made at both time points. It is possible that elements involved in the somatic yoga intervention within this study, including breathing techniques and sensory-awareness exercises, help to direct and adaptively train attention and response inhibition, cognitive abilities that have previously been shown to be implicated in FND (Alluri et al., 2020; Millman et al., 2025; Pick et al., 2019; Pick et al., 2026; Van Patten et al., 2024). It is also a possibility that the elevations in noticing and body listening that were self-reported in this yoga sample at week 3 and/or week 6 (Kennedy-Barnes et al., 2026) could be related to this improved cognitive control, something that should be explored in a larger-scale trial.

A review by Voss, Cerna & Gothe (Voss et al., 2023) put forward improved stress regulation and improved neurocognitive resource efficiency as two mechanisms that may contribute to the cognitive improvements seen in yoga. In line with this, after a single session of yoga or music, self-reported state negative affect decreased, suggesting positive effects of both interventions on current mood. Further, after a single session and post-six weeks, state dissociative symptoms decreased while positive affect increased in both groups, with a larger magnitude of change in the yoga group. The relaxation, regulative breathing, and restorative postures included in the somatic yoga intervention may have helped activate the parasympathetic nervous system, support autonomic and/or emotional regulation, and enhance present state grounding and positive mood (Field, 2016; Gothe et al., 2019; Streeter et al., 2012). Mood shifts in the form of lower depression scores (at six weeks) were similarly seen in this yoga sample (Kennedy-Barnes et al., 2026), possibly reflective of a cumulative effect of state changes that may have taken place across the weeks of practice. These findings align with other explorations of yoga and mindfulness/body-based practices, including significant improvements in overall present state awareness after a Mindfulness and Movements of Integration (MMI) program (Bloise et al., 2016), more self-reported frequent use of cognitive reappraisal in long-term yoga practitioners (Kobylińska et al., 2018), elevated self-reported emotion regulation in adolescents practicing yoga relative to non-practicing adolescents (Janjhua et al., 2020), and improved stress reactivity and recovery (blood pressure, salivary cortisol) (Benvenutti et al., 2017) or anti-inflammatory effects (Grzenda et al., 2024) after yoga. Yoga may encourage an integration of both top-down and bottom-up processes (Gard et al., 2014), promoting a bidirectional interaction of mind and body (Voss et al., 2023).

The suggestion of improved stress regulation being a mechanism at play in the practice of yoga is further corroborated by the decrease in autonomic arousal (average heartrate) seen in both the yoga and music groups immediately after a single session. This result points towards potential positive relaxation effects of both types of intervention in this population and adds to the evidence base for the positive impact of yoga and music on parasympathetic activation and stress reduction (de Witte et al., 2022; Streeter et al., 2012; Zaccaro et al., 2018). Given converging evidence for elevated autonomic arousal and hyper-reactivity in FND (Pick et al., 2019), interventions which help to target these processes and promote improved regulation of bodily signals may be particularly beneficial due to links between autonomic regulation, affective states, and symptomatology. Although changes in objective autonomic arousal were not seen within the emotion regulation task, both participant groups reported elevated subjective arousal and less pleasant valence ratings when viewing negative images, regardless of timepoint.

No changes were seen regarding state cardiac interoception measures (heartbeat tracking accuracy or confidence) in the laboratory. This contrasts with improvement in aspects of interoceptive sensibility seen in this sample (Kennedy-Barnes et al., 2026), and underscores evidence for altered trait interoceptive sensibility possibly being more central to FND (Millman et al., 2023; Pick et al., 2020). It is also possible that state interoceptive changes which may have occurred due to yoga were not picked up in the cardiac domain, with other domains of state interoceptive accuracy, including respiratory or gastric (Suksasilp & Garfinkel, 2022), worth measurement in future research.

As detailed in Kennedy-Barnes et al. (Kennedy-Barnes et al., 2026), this study is the first feasibility randomised controlled trial to evaluate yoga in adults with FND. Given the aims and nature of the trial, the sample size is acceptable but not powered for hypothesis testing, and larger studies are needed to evaluate efficacy and test hypotheses regarding mechanisms of action. With regards to intervention delivery, this occurred over a relatively short-term, six-week period, which may not be sufficient to capture longer-term changes. It would also be worth incorporating extended follow-ups (beyond the 3-months reported in Kennedy-Barnes et al., 2026), to evaluate possible sustained or evolving effects of the intervention. While the trial involved one-to-one sessions which allowed for individual tailoring and support, there could also be benefits to group sessions which would be worth exploring in future research.

The music-based control condition seemed to involve relaxation and parasympathetic enhancement, making it difficult to fully isolate effects specific to somatic yoga. Future trials could benefit from the inclusion of an alternative active control condition that matches non-specific factors, like therapist contact, without mechanistic overlap. Further, although participants were asked to abstain from taking psychotropic medications prior to the laboratory visits if possible to do so, the possible effects of these types of medications, particularly regarding cognitive task performance, warrant future consideration. Additionally, given reports of elevated subjective cognitive symptoms in FND (Pick et al., 2023, 2026), and significant, large effects of yoga training on subjective cognitive impairment (Grzenda et al., 2024), it may be worth measuring these types of symptoms alongside objective cognitive functioning in future studies.

## Conclusions

This study provides further evidence for the use of yoga for FND (Kennedy-Barnes et al., 2026; Kipnis et al., 2024; Park et al., 2021), adding to a growing evidence base for the use of mind-body interventions in this population. Although preliminary, the results are suggestive of potential mechanisms of action, including cognitive control, autonomic arousal, and affective processing, all of which may play varying roles in the experience of FND symptoms. These findings warrant future, larger-scale, controlled trials to evaluate efficacy and mechanisms.

## Data Availability

All data produced in the present study are available upon reasonable request to the authors

## Acknowledgements

We are grateful to all participants and those who contributed to the patient and public involvement input on this study. With thanks also to FND Hope UK for assistance with recruitment.

## CRediT statement

Conceptualisation – SP, EKB.

Methodology – SP, EKB, LSMM.

Validation – SP, JH.

Formal analysis – LSMM, JH.

Investigation (data collection/recruitment) – LSMM, SP, EKB, YB, AD, JP.

Resources – SP.

Data curation (data processing/preparation) – LSMM, EKB, YB, SP, AD, JP.

Writing (original draft) – LSMM.

Writing (review/editing) – SP, EKB, YB.

Supervision – SP, LSMM.

Project administration – SP, EKB, LSMM.

Funding acquisition – SP.

