## Supplementary Materials for "Somatic yoga therapy for functional neurological disorder: An experimental pilot study examining cognitive and affective mechanisms"

**Table S1.** Standardised instructions – heartbeat tracking task

| **Instructions 1** | In this heartbeat tracking task you will be asked to count your heartbeat for several periods of time.  Please do not attempt to take your pulse or use any other physical aid to count your heartbeats.  Do not try to count the number of seconds that pass during the heartbeat tracking periods.  Press space to continue. |
| --- | --- |
| **Instructions 2** | At the start of each heartbeat tracking period, you will be prompted with the following words on screen:  ‘START COUNTING YOUR HEARTBEATS’  When the instruction appears, please concentrate fully on your heartbeat and try to count each one, until you see the words:  ‘STOP COUNTING’  Press space to continue. |
| **Instructions 3** | Immediately after each heartbeat tracking period, you will be asked for two judgements.  First, you will be asked how many heartbeats you counted.  Please only count heartbeats that you can genuinely feel, and answer ‘0’ if you cannot feel any heartbeats at all.  Please give your answer using the number keys on the keyboard, followed by ‘space’ to move on.  Press space to continue. |
| **Instructions 4** | Next, you will be asked how confident you were in your first answer from 0-10.  On this scale, 0 = Not at all confident and 10 = Very confident.  You can choose any number from 0-10.  After you have entered your response, you can move on by pressing space.  Press space to continue. |
| **Instructions 5** | Each heartbeat tracking period will be separated by a rest period, when the word ‘Rest’ will be shown on screen.  During the rest period, please sit quietly and relax, but do not try to track your heartbeat or take your pulse.  Please try to keep your eyes on the screen and stay as still as you can throughout the task.  Press space to continue |
| **Instructions 6** | You can now practise the heartbeat tracking task.  Press space to continue. |
|  | **PRACTISE TRIAL (20s)** |
| **Instructions 7** | You have now completed the practice heartbeat tracking period.  Feel free to ask any questions now.  It is very important that you stay as still as you possibly can during this task.  Press space when you are ready to start the task. |

**Table S2.** Standardised instructions – affective images task

| **Instructions 1** | In this task, you will be shown lots of different pictures on the screen. The pictures will be shown in sets of four, presented one after another.  There will be 12 sets of pictures, separated by brief breaks, when you will see a cross in the centre of the screen.  During the breaks, please try to relax and stay focused on the cross.  Please press space to continue. |
| --- | --- |
| **Instructions 2** | Before some of the blocks of pictures, you will see the word ‘Watch’ on screen for a few seconds.  When you see this, please keep your eyes on the screen and just look at the subsequent pictures.  Do not try to change how you feel when you view the pictures.  Please press space to continue. |
| **Instructions 3** | Before the other blocks of pictures, you will see the word ‘Dampen’ on screen.  When you see this, please try to minimise your reactions to the pictures. You can use any strategy – the important thing is that you try to reduce the intensity of your emotional response to the pictures.  Please press space to continue. |
| **Instructions 4** | After every set of pictures, you will be asked two simple questions about how you feel, right then, in that moment.  One rating is about whether you are experiencing pleasant or unpleasant feelings.  The scale goes from unpleasant-pleasant, from 1-9.  If you felt completely unhappy (pleasant, satisfied, contented, hopeful), press 9.  If you felt completely unhappy (annoyed, unsatisfied, bored), press 1.  You can also describe feelings in between, by pressing any number from 1 to 9.  If you felt completely neutral, select 5.  Press space to see an example of the scale. |
|  | **EXAMPLE VALENCE SCALE** |
| **Instructions 5** | The other rating is about whether you feel calm or excited.  The scale goes from feeling completely relaxed (sluggish, sleepy, calm) to feeling completely excited (stimulated, jittery, wide awake, aroused).  If you felt completely aroused or excited press 9.  If you felt totally relaxed and calm press 1.  Again, you can describe feelings in between by pressing any number from 1-9.  If you did not feel at all excited or at all calm you would select 5.  Press space to see an example of the scale. |
|  | **EXAMPLE AROUSAL SCALE** |
| **Instructions 6** | Some of the pictures may make you feel strong emotions, others might not affect you at all.  Please rate your immediate personal experience after each set of pictures.  You will have 5 seconds to respond – when you have entered your response, please wait for the task to move on.  You will not always be asked to make your ratings in the same order.  You will now see some example pictures, so that you can practice the task.  Please press space to begin the practice. |
|  | **PRACTISE TRIALS** |
| **Instructions 7** | You have now completed the practice items.  Please remember to look at the screen and stay as still as possible during this task.  Remember, when you see the word ‘Watch’, just look at the pictures.  When you see the word ‘Dampen’, try to minimise your emotional reactions to the images.  Feel free to ask any questions now and let the researcher know that you are ready to start.  Please press space to begin. |

**Table S3.** Raw descriptive statistics for all laboratory measures.

|  | **Yoga** | | | **Music** | | |
| --- | --- | --- | --- | --- | --- | --- |
|  | **Baseline**  M (SD) | **Post single session**  M (SD) | **Post 6 weeks**  M (SD) | **Baseline**    M (SD) | **Post single session**  M (SD) | **Post 6 weeks**  M (SD) |
| **HTT Accuracy** | .39 (3.17) | .49 (.34) | .53 (.35) | .32 (.29) | .44 (.39) | .42 (.35) |
| **HTT Confidence** | 3.90 (3.17) | 2.88 (3.05) | 4.48 (3.19) | 5.70 (3.09) | 5.91 (3.29) | 6.28 (3.20) |
| **RVPMDL** | 544 (148) | 457 (58.50) | 465 (81.70) | 483 (81.50) | 461 (58.00) | 499 (127) |
| **RVPA** | .87 (.05) | .90 (.05) | .92 (.06) | .91 (.04) | .93 (.04) | .93 (.03) |
| **RVPPH** | .50 (.20) | .63 (.18) | .71 (.22) | .66 (.15) | .74 (.17) | .74 (.13) |
| **RVPPFA** | .01 (.01) | .01 (.01) | .01 (.01) | .01 (.01) | .00 (.01) | .00 (.01) |
| **RVPTM** | 27 (10.60) | 20 (9.90) | 15.80 (12.10) | 18.30 (7.98) | 14.20 (9.41) | 14.20 (6.76) |
| **SSTSSRT** | 276 (62.90) | 249 (50.40) | 264 (53.80) | 231 (28.90) | 220 (29.40) | 224 (26.30) |
| **SSTDEG** | 2.42 (3.03) | 1.91 (1.64) | 1.00 (1.41) | .90 (1.45) | 1.12 (1.13) | 1.25 (1.98) |
| **SSTDES** | 41.70 (3.06) | 41.40 (4.48) | 41.50 (3.27) | 36.20 (3.62) | 35.40 (6.09) | 38.80 (1.91) |
| **SSTMT** | 7.50 (4.94) | 4.36 (5.71) | 3.09 (3.27) | 15.90 (24.20) | 5.12 (8.15) | 5.62 (7.31) |
| **CADSS** | 11.00 (8.82) | 6.82 (8.42) | 6.82 (7.08) | 4.09 (4.32) | 4.22 (5.87) | 1.71 (2.29) |
| **Positive Affect** | 23.60 (8.69) | 33.90 (11.50) | 34.00 (7.17) | 25.30 (9.35) | 30.40 (7.94) | 30.70 (7.99) |
| **Negative Affect** | 16.10 (4.78) | 11.80 (2.82) | 15.40 (5.56) | 14.60 (4.36) | 11.90 (2.53) | 13.60 (3.64) |
| **Baseline Heartrate** | 74.60 (8.29) | 69.20 (8.34) | 71.50 (11.00) | 73.80 (9.06) | 64.60 (9.82) | 73.00 (15.60) |
| **Baseline EDA** | 10.20 (4.96) | 9.78 (2.92) | 7.66 (2.09) | 13.10 (11.40) | 10.60 (6.19) | 9.66 (4.43) |
| **Baseline RMSSD** | 65.00 (59.60) | 72.80 (67.20) | 77.50 (81.10) | 37.00 (41.90) | 58.20 (50.30) | 40.20 (45.60) |
| **Arousal** |  |  |  |  |  |  |
| *NeutralWatch* | 4.24 (1.70) | 4.15 (1.46) | 3.78 (1.51) | 3.55 (1.66) | 3.09 (1.53) | 3.87 (1.94) |
| *NegativeWatch* | 5.67 (1.79) | 5.19 (1.66) | 5.20 (2.15) | 5.24 (2.16) | 4.33 (1.82) | 5.04 (1.75) |
| *NegativeDampen* | 5.67 (1.25) | 5.24 (1.39) | 4.58 (1.55) | 5.24 (2.16) | 4.40 (1.68) | 5.13 (1.80) |
| **Valence** |  |  |  |  |  |  |
| *NeutralWatch* | 5.09 (1.23) | 5.31 (.76) | 5.45 (1.18) | 5.02 (1.21) | 5.42 (1.84) | 5.48 (1.03) |
| *NegativeWatch* | 3.74 (1.85) | 4.24 (1.15) | 3.58 (1.62) | 3.76 (1.72) | 4.36 (1.66) | 4.14 (1.46) |
| *NegativeDampen* | 3.68 (1.27) | 4.31 (1.21) | 4.23 (1.42) | 3.49 (1.61) | 4.21 (1.92) | 4.09 (1.47) |
| **Heartrate** |  |  |  |  |  |  |
| *NeutralWatch* | 70.30 (8.49) | 66.20 (7.78) | 68.40 (10.80) | 68.60 (8.20) | 64.60 (10.00) | 68.60 (11.20) |
| *NegativeWatch* | 69.60 (8.07) | 65.50 (8.07) | 67.90 (10.70) | 68.80 (8.11) | 64.20 (10.00) | 67.40 (11.60) |
| *NegativeDampen* | 69.70 (7.93) | 66.00 (7.76) | 66.30 (10.10) | 68.60 (8.69) | 64.60 (10.10) | 67.60 (11.00) |
| **EDA** |  |  |  |  |  |  |
| *NeutralWatch* | 11.00 (3.81) | 8.80 (2.82) | 7.78 (2.04) | 15.90 (12.20) | 13.00 (7.73) | 9.80 (4.40) |
| *NegativeWatch* | 10.90 (3.48) | 8.97 (2.88) | 7.68 (2.31) | 16.00 (12.10) | 13.00 (7.88) | 9.83 (4.43) |
| *NegativeDampen* | 10.70 (3.54) | 9.31 (3.18) | 7.75 (2.11) | 15.70 (12.00) | 13.00 (7.91) | 9.62 (4.37) |
| **RMSSD** |  |  |  |  |  |  |
| *NeutralWatch* | 42.80 (44.40) | 59.60 (73.20) | 50.90 (64.90) | 80.20 (132.00) | 31.70 (29.10) | 20.50 (14.20) |
| *NegativeWatch* | 45.00 (47.30) | 54.80 (69.60) | 60.70 (88.70) | 43.20 (81.40) | 28.90 (15.80) | 24.90 (20.30) |
| *NegativeDampen* | 61.20 (89.30) | 109.00 (131.00) | 67.60 (100.00) | 29.00 (22.80) | 49.00 (72.30) | 41.10 (72.30) |

*Notes.* HTT = heartbeat tracking task; RVP = rapid visual information processing task; RVPMDL = RVP median response latency; RVPA = RVP sensitivity to target sequence; RVPPH = RVP probability of hit; RVPPFA = RVP probability of false alarm; RVPTM = RVP total misses; SST = stop signal task; SSTSSRT = stop signal reaction time; SSTDEG = SST direction errors go trials; SSTDES = SST direction errors stop trials; SSTMT = SST missed trials; CADSS = clinician administered dissociative states scale; EDA = electrodermal activity; RMSSD = root mean square of successive differences.

**Table S4.** Linear mixed effects models pre-post single session. – Add reference to main manuscript

|  | ***ß*** | **SE** | **95% CI** | **t** | **p** |
| --- | --- | --- | --- | --- | --- |
| **HTT Accuracy** |  |  |  |  |  |
| *Intercept* | .41 | .34 | -.23, 1.05 | 1.22 | .23 |
| *Group* | -.12 | .22 | -.54, .30 | -.54 | .59 |
| *Time* | .05 | .19 | -.31, .42 | .28 | .79 |
| *Group x Time* | .05 | .12 | -.20, .29 | .38 | .71 |
| **HTT Confidence** |  |  |  |  |  |
| *Intercept* | 3.94 | 3.55 | -2.86, 10.80 | 1.11 | .28 |
| *Group* | .98 | 2.31 | -3.50, 5.39 | .43 | .67 |
| *Time* | -1.85 | 2.08 | -5.98, 2.17 | -.89 | .39 |
| *Group x Time* | .83 | 1.39 | -1.84, 3.60 | .60 | .56 |
| **RVPMDL** |  |  |  |  |  |
| *Intercept* | 756.83 | 133.85 | 498.76, 1014.59 | 5.65 | .00 |
| *Group* | -126.61 | 84.66 | -289.54, 36.89 | -1.50 | .15 |
| *Time* | -152.62 | 81.76 | -311.34, 6.85 | -1.87 | .08 |
| *Group x Time* | 66.05 | 53.14 | -37.73, 168.93 | 1.24 | .23 |
| **RVPA** |  |  |  |  |  |
| *Intercept* | .76 | .04 | .68, .84 | 18.41 | .00 |
| *Group* | .07 | .03 | .02, .12 | 2.55 | .02 |
| *Time* | .06 | .02 | .02, .10 | 2.95 | .01 |
| *Group x Time* | -.02 | .01 | -.04, .01 | -1.38 | .19 |
| **RVPPH** |  |  |  |  |  |
| *Intercept* | .03 | .16 | -.27, .34 | .20 | .84 |
| *Group* | .27 | .10 | .08, .47 | 2.71 | .01 |
| *Time* | .24 | .08 | .09, .39 | 3.18 | .01 |
| *Group x Time* | -.08 | .05 | -.18, .02 | -1.63 | .12 |
| **RVPPFA** |  |  |  |  |  |
| *Intercept* | .00 | .01 | -.02, .02 | .22 | .83 |
| *Group* | .00 | .01 | -.01, .02 | .64 | .53 |
| *Time* | .00 | .01 | -.01, .02 | .58 | .57 |
| *Group x Time* | -.00 | .00 | -.01, .00 | -.90 | .38 |
| **RVPTM** |  |  |  |  |  |
| *Intercept* | 52.26 | 8.63 | 35.74, 68.74 | 6.06 | .00 |
| *Group* | -14.84 | 5.47 | -25.27, -4.39 | -2.71 | .01 |
| *Time* | -13.11 | 4.12 | -21.04, -4.92 | -3.18 | .01 |
| *Group x Time* | 4.40 | 2.69 | -.91, 9.60 | 1.63 | .12 |
| **SSTSSRT** |  |  |  |  |  |
| *Intercept* | 356.05 | 54.38 | 252.16, 461.05 | 6.55 | .00 |
| *Group* | -55.06 | 35.58 | -123.84, 12.90 | -1.55 | .13 |
| *Time* | -34.87 | 32.33 | -99.03, 27.33 | -1.08 | .30 |
| *Group x Time* | 10.01 | 21.39 | -31.07, 52.52 | .47 | .65 |
| **SSTDEG** |  |  |  |  |  |
| *Intercept* | 5.17 | 3.06 | -.65, 11.00 | 1.69 | .10 |
| *Group* | -2.25 | 2.00 | -6.06, 1.56 | -1.13 | .27 |
| *Time* | -1.24 | 1.97 | -4.99, 2.51 | -.63 | .53 |
| *Group x Time* | .73 | 1.30 | -1.74, 3.20 | .57 | .58 |
| **SSTDES** |  |  |  |  |  |
| *Intercept* | 47.64 | 4.26 | 39.48, 55.77 | 11.18 | .00 |
| *Group* | -5.50 | 2.79 | -10.82, -.16 | -1.97 | .06 |
| *Time* | -.51 | 2.33 | -5.01, 4.10 | -.22 | .83 |
| *Group x Time* | .04 | 1.55 | -3.02, 3.02 | .02 | .98 |
| **SSTMT** |  |  |  |  |  |
| *Intercept* | -6.32 | 17.86 | -40.55, 28.57 | -.35 | .73 |
| *Group* | 16.85 | 11.68 | -6.07, 39.20 | 1.44 | .16 |
| *Time* | 5.42 | 11.27 | -17.06, 27.07 | .48 | .64 |
| *Group x Time* | -8.45 | 7.43 | -22.67, 6.49 | -1.14 | .27 |
| **CADSS** |  |  |  |  |  |
| *Intercept* | 27.46 | 6.62 | 14.81, 40.10 | 4.15 | .00 |
| *Group* | -11.66 | 4.26 | -19.79, -3.52 | -2.74 | .01 |
| *Time* | -9.55 | 3.40 | -16.14, -2.86 | -2.81 | .01 |
| *Group x Time* | 4.75 | 2.22 | .41, 9.05 | 2.15 | .05 |
| **Positive Affect** |  |  |  |  |  |
| *Intercept* | 4.20 | 8.68 | -12.37, 20.78 | .48 | .63 |
| *Group* | 9.05 | 5.63 | -1.71, 19.80 | 1.61 | .12 |
| *Time* | 17.69 | 4.31 | 9.17, 26.01 | 4.11 | .00 |
| *Group x Time* | -7.36 | 2.89 | -12.92, -1.60 | -2.55 | .02 |
| **Negative Affect** |  |  |  |  |  |
| *Intercept* | 23.79 | 4.13 | 15.83, 31.68 | 5.76 | .00 |
| *Group* | -3.45 | 2.68 | -8.58, 1.73 | -1.29 | .21 |
| *Time* | -6.26 | 2.36 | -10.81, -1.58 | -2.65 | .02 |
| *Group x Time* | 2.01 | 1.58 | -1.13, 5.03 | 1.27 | .22 |
| **Baseline Heartrate** |  |  |  |  |  |
| *Intercept* | 78.20 | 6.68 | 65.40, 91.01 | 11.71 | .00 |
| *Group* | 1.73 | 4.30 | -6.52, 9.96 | .40 | .69 |
| *Time* | -2.86 | 2.48 | -7.68, 2.01 | -1.15 | .26 |
| *Group x Time* | -2.51 | 1.64 | -5.73, .66 | -1.54 | .14 |
| **Baseline EDA** |  |  |  |  |  |
| *Intercept* | 5.262 | 6.54 | -7.19, 17.73 | .81 | .43 |
| *Group* | 5.15 | 4.31 | -3.07, 13.35 | 1.20 | .24 |
| *Time* | 2.31 | 3.48 | -4.50, 9.05 | .67 | .52 |
| *Group x Time* | -2.53 | 2.28 | -6.93, 1.92 | -1.11 | .28 |
| **Baseline RMSSD** |  |  |  |  |  |
| *Intercept* | 99.31 | 67.87 | -31.52, 229.91 | 1.46 | .15 |
| *Group* | -42.24 | 43.86 | -126.59, 42.33 | -.96 | .34 |
| *Time* | -6.39 | 40.77 | -85.76, 73.34 | -.16 | .88 |
| *Group x Time* | 14.28 | 26.73 | -37.99, 66.20 | .53 | .60 |
| **Arousal** |  |  |  |  |  |
| *Intercept* | 3.54 | .35 | 2.87, 4.21 | 10.19 | .00 |
| *Group: Yoga* | .71 | .48 | -.21, 1.64 | 1.48 | .15 |
| *Time: Post* | -.50 | .35 | -1.18, .18 | -1.42 | .16 |
| *Condition: NegativeWatch* | 1.76 | .33 | 1.12, 2.41 | 5.32 | .00 |
| *Condition: NegativeDampen* | 1.69 | .33 | 1.04, 2.33 | 5.08 | .00 |
| *Group x Time: Yoga x Post* | .37 | .49 | -.58, 1.32 | .77 | .45 |
| *GroupxCondition: Yoga x NegativeWatch* | -.40 | .46 | -1.28, .50 | -.86 | .39 |
| *Group x Condition: Yoga x NegativeDampen* | -.28 | .45 | -1.16, .60 | -.62 | .54 |
| *Time x Condition: Post x NegativeWatch* | -.54 | .49 | -1.49, .41 | -1.11 | .27 |
| *Time x Condition: Post x NegativeDampen* | -.41 | .49 | -1.36, .54 | -.84 | .40 |
| *Group x Time x Condition: Yoga x Post x NegativeWatch* | .19 | .69 | -1.15, 1.52 | .27 | .78 |
| *Group x Time x Condition: Yoga x Post x NegativeDampen* | .04 | .68 | -1.28, 1.37 | .07 | .95 |
| **Valence** |  |  |  |  |  |
| *Intercept* | 5.00 | .28 | 4.45, 5.53 | 17.80 | .00 |
| *Group: Yoga* | .11 | .39 | -.64, .87 | .29 | .77 |
| *Time: Post* | .43 | .32 | -.19, 1.04 | 1.36 | .18 |
| *Condition: NegativeWatch* | -1.23 | .30 | -1.82, -.64 | -4.08 | .00 |
| *Condition: NegativeDampen* | -1.51 | .30 | -2.10, .-.93 | -5.06 | .00 |
| *Group x Time: Yoga x Post* | -.17 | .45 | -1.04, .70 | -.39 | .70 |
| *GroupxCondition: Yoga x NegativeWatch* | -.17 | .42 | -.99, .65 | -.41 | .68 |
| *Group x Condition: Yoga x NegativeDampen* | .09 | .42 | -.72, .91 | .21 | .83 |
| *Time x Condition: Post x NegativeWatch* | .10 | .45 | -.77, .97 | .22 | .83 |
| *Time x Condition: Post x NegativeDampen* | .33 | .44 | -.54, 1.19 | .73 | .46 |
| *Group x Time x Condition: Yoga x Post x NegativeWatch* | .23 | .63 | -1.01, 1.45 | .36 | .72 |
| *Group x Time x Condition: Yoga x Post x NegativeDampen* | .12 | .63 | -1.10, 1.33 | .19 | .85 |
| **Heartrate** |  |  |  |  |  |
| *Intercept* | 68.60 | 2.57 | 63.58, 73.62 | 26.67 | .00 |
| *Group: Yoga* | 1.72 | 3.56 | -5.23, 8.67 | .48 | .63 |
| *Time: Post* | -2.82 | .64 | -4.07, -1.58 | -4.39 | .00 |
| *Condition: NegativeWatch* | .20 | .60 | -.96, 1.37 | .34 | .73 |
| *Condition: NegativeDampen* | -.01 | .60 | -1.17, 1.16 | -.01 | .99 |
| *Group x Time: Yoga x Post* | 1.15 | .91 | -.62, 2.92 | 1.26 | .21 |
| *GroupxCondition: Yoga x NegativeWatch* | -1.02 | .83 | -2.63, .60 | -1.22 | .22 |
| *Group x Condition: Yoga x NegativeDampen* | -.73 | .83 | -2.35, .88 | -.88 | .38 |
| *Time x Condition: Post x NegativeWatch* | -.57 | .89 | -2.31, 1.16 | -.64 | .52 |
| *Time x Condition: Post x NegativeDampen* | -.01 | .89 | -1.75, 1.72 | -.01 | .99 |
| *Group x Time x Condition: Yoga x Post x NegativeWatch* | .71 | 1.26 | -1.75, 3.16 | .56 | .58 |
| *Group x Time x Condition: Yoga x Post x NegativeDampen* | .57 | 1.26 | -1.89, 3.02 | .45 | .65 |
| **EDA** |  |  |  |  |  |
| *Intercept* | 15.83 | 2.19 | 11.55, 20.11 | 7.21 | .00 |
| *Group: Yoga* | -4.81 | 3.04 | -10.74, 1.11 | -1.59 | .13 |
| *Time: Post* | -4.76 | .55 | -5.82, -3.69 | -8.67 | .00 |
| *Condition: NegativeWatch* | .17 | .52 | -.84, 1.18 | .33 | .74 |
| *Condition: NegativeDampen* | -.25 | .52 | -1.27, .76 | -.48 | .63 |
| *Group x Time: Yoga x Post* | 2.64 | .77 | 1.14, 4.13 | 3.42 | .00 |
| *GroupxCondition: Yoga x NegativeWatch* | -.06 | .71 | -1.44, 1.31 | -.09 | .93 |
| *Group x Condition: Yoga x NegativeDampen* | .14 | .71 | -1.24, 1.51 | .19 | .85 |
| *Time x Condition: Post x NegativeWatch* | -.10 | .75 | -1.56, 1.37 | -.13 | .90 |
| *Time x Condition: Post x NegativeDampen* | .28 | .76 | -1.18, 1.75 | .37 | .71 |
| *Group x Time x Condition: Yoga x Post x NegativeWatch* | -.16 | 1.06 | -2.22, 1.89 | -.15 | .88 |
| *Group x Time x Condition: Yoga x Post x NegativeDampen* | .01 | 1.06 | -2.05, 2.07 | .01 | .99 |
| **RMSSD** |  |  |  |  |  |
| *Intercept* | 43.99 | 12.57 | 19.79, 68.21 | 3.50 | .00 |
| *Group: Yoga* | 20.88 | 17.23 | -12.31, 54.05 | 1.21 | .23 |
| *Time: Post* | 4.19 | 17.20 | -29.24, 37.60 | .24 | .81 |
| *Condition: NegativeWatch* | 4.08 | 16.55 | -28.09, 36.22 | .25 | .81 |
| *Condition: NegativeDampen* | -7.05 | 16.55 | -39.22, 25.10 | -.43 | .67 |
| *Group x Time: Yoga x Post* | 14.45 | 25.96 | -35.49, 65.79 | .56 | .58 |
| *GroupxCondition: Yoga x NegativeWatch* | -5.31 | 22.78 | -49.56, 38.97 | -.23 | .82 |
| *Group x Condition: Yoga x NegativeDampen* | -16.39 | 22.78 | -60.64, 27.89 | -.72 | .47 |
| *Time x Condition: Post x NegativeWatch* | 24.44 | 24.81 | -23.61, 72.78 | .99 | .33 |
| *Time x Condition: Post x NegativeDampen* | 5.11 | 24.81 | -43.41, 53.05 | .21 | .84 |
| *Group x Time x Condition: Yoga x Post x NegativeWatch* | -55.16 | 35.72 | -124.96, 13.88 | -1.54 | .12 |
| *Group x Time x Condition: Yoga x Post x NegativeDampen* | -29.31 | 35.19 | -98.11, 38.67 | -.83 | .41 |

*Notes.* HTT = heartbeat tracking task; RVP = rapid visual information processing task; RVPMDL = RVP median response latency; RVPA = RVP sensitivity to target sequence; RVPPH = RVP probability of hit; RVPPFA = RVP probability of false alarm; RVPTM = RVP total misses; SST = stop signal task; SSTSSRT = stop signal reaction time; SSTDEG = SST direction errors go trials; SSTDES = SST direction errors stop trials; SSTMT = SST missed trials; CADSS = clinician administered dissociative states scale; EDA = electrodermal activity; RMSSD = root mean square of successive differences.

**Table S5.** Missing data across all laboratory measures.

|  | **Yoga** | | | **Music** | | |
| --- | --- | --- | --- | --- | --- | --- |
|  | **Baseline**  n (%) | **Post single session**  n (%) | **Post 6 weeks**  n (%) | **Baseline**    n (%) | **Post single session**  n (%) | **Post 6 weeks**  n (%) |
| **HTT Accuracy** | 0 (0) | 0 (0) | 1 (8) | 0 (0) | 3 (27) | 3 (27) |
| **HTT Confidence** | 0 (0) | 0 (0) | 1 (8) | 0 (0) | 3 (27) | 3 (27) |
| **RVPMDL** | 2 (16) | 0 (0) | 1 (8) | 0 (0) | 3 (27) | 3 (27) |
| **RVPA** | 2 (16) | 0 (0) | 1 (8) | 0 (0) | 3 (27) | 3 (27) |
| **RVPPH** | 2 (16) | 0 (0) | 1 (8) | 0 (0) | 3 (27) | 3 (27) |
| **RVPPFA** | 2 (16) | 0 (0) | 1 (8) | 0 (0) | 3 (27) | 3 (27) |
| **RVPTM** | 2 (16) | 0 (0) | 1 (8) | 0 (0) | 3 (27) | 3 (27) |
| **SSTSSRT** | 0 (0) | 1 (8) | 1 (8) | 1 (9) | 3 (27) | 3 (27) |
| **SSTDEG** | 0 (0) | 1 (8) | 1 (8) | 1 (9) | 3 (27) | 3 (27) |
| **SSTDES** | 0 (0) | 1 (8) | 1 (8) | 1 (9) | 3 (27) | 3 (27) |
| **SSTMT** | 0 (0) | 1 (8) | 1 (8) | 1 (9) | 3 (27) | 3 (27) |
| **CADSS** | 0 (0) | 0 (0) | 1 (8) | 0 (0) | 2 (18) | 4 (36) |
| **Positive Affect** | 0 (0) | 0 (0) | 1 (8) | 0 (0) | 3 (27) | 4 (36) |
| **Negative Affect** | 0 (0) | 0 (0) | 1 (8) | 0 (0) | 3 (27) | 4 (36) |
| **Baseline Heartrate** | 0 (0) | 0 (0) | 1 (8) | 0 (0) | 2 (18) | 3 (27) |
| **Baseline EDA** | 0 (0) | 2 (17) | 1 (8) | 2 (18) | 2 (18) | 3 (27) |
| **Baseline RMSSD** | 0 (0) | 0 (0) | 1 (8) | 0 (0) | 2 (18) | 3 (27) |
| **Arousal** |  |  |  |  |  |  |
| *NeutralWatch* | 2 (4) | 14 (29) | 6 (13) | 2 (5) | 11 (25) | 13 (30) |
| *NegativeWatch* | 6 (13) | 16 (33) | 8 (17) | 6 (14) | 8 (18) | 16 (36) |
| *NegativeDampen* | 2 (4) | 14 (29) | 7 (15) | 6 (14) | 9 (20) | 14 (32) |
| **Valence** |  |  |  |  |  |  |
| *NeutralWatch* | 5 (10) | 13 (27) | 8 (17) | 3 (7) | 8 (18) | 13 (30) |
| *NegativeWatch* | 6 (13) | 15 (31) | 10 (21) | 2 (5) | 11 (25) | 16 (36) |
| *NegativeDampen* | 4 (8) | 13 (27) | 9 (19) | 1 (2) | 10 (23) | 12 (27) |
| **Heartrate** |  |  |  |  |  |  |
| *NeutralWatch* | 0 (0) | 14 (29) | 10 (21) | 0 (0) | 8 (18) | 13 (30) |
| *NegativeWatch* | 1 (2) | 14 (29) | 8 (17) | 0 (0) | 8 (18) | 14 (32) |
| *NegativeDampen* | 1 (2) | 14 (29) | 7 (15) | 0 (0) | 8 (18) | 13 (30) |
| **EDA** |  |  |  |  |  |  |
| *NeutralWatch* | 1 (2) | 16 (33) | 10 (21) | 5 (11) | 9 (20) | 13 (30) |
| *NegativeWatch* | 3 (6) | 15 (31) | 8 (17) | 5 (11) | 8 (18) | 14 (32) |
| *NegativeDampen* | 3 (6) | 15 (31) | 7 (15) | 6 (14) | 8 (18) | 13 (30) |
| **RMSSD** |  |  |  |  |  |  |
| *NeutralWatch* | 0 (0) | 14 (29) | 10 (21) | 0 (0) | 8 (18) | 13 (30) |
| *NegativeWatch* | 1 (2) | 14 (29) | 8 (17) | 0 (0) | 8 (18) | 14 (32) |
| *NegativeDampen* | 1 (2) | 14 (29) | 7 (15) | 0 (0) | 8 (18) | 13 (30) |

*Notes.* HTT = heartbeat tracking task; RVP = rapid visual information processing task; RVPMDL = RVP median response latency; RVPA = RVP sensitivity to target sequence; RVPPH = RVP probability of hit; RVPPFA = RVP probability of false alarm; RVPTM = RVP total misses; SST = stop signal task; SSTSSRT = stop signal reaction time; SSTDEG = SST direction errors go trials; SSTDES = SST direction errors stop trials; SSTMT = SST missed trials; CADSS = clinician administered dissociative states scale; EDA = electrodermal activity; RMSSD = root mean square of successive differences.

**Table S6.** Three-way mixed ANOVAs pre-post single session.

| **Variable** | **ANOVA** | **F (df)** | ***p*** | ***ηp2*** |
| --- | --- | --- | --- | --- |
| Arousal | Group x Time x Condition | .04 (2, 422.59) | .96 | .00 |
| Valence | Group x Time x Condition | .06 (2, 429.38) | .94 | .00 |
| Heart rate | Group x Time x Condition | .18 (2, 451.00) | .84 | .00 |
| Skin conductance | Group x Time x Condition | .02 (2, 425.03) | .98 | .00 |
| RMSSD | Group x Time x Condition | 1.03 (2, 451.93) | .36 | .00 |

*Notes.* RMSSD = root mean square of successive differences.

**Table S7.** Laboratory measures by group across time post-single session (emmeans).

|  | **Yoga** |  | **Music** |  |
| --- | --- | --- | --- | --- |
|  | **Baseline** | **Post single session** | **Baseline** | **Post single session** |
| **HTT Accuracy** | .39 | .49 | .32 | .47 |
| **HTT Confidence** | 3.90 | 2.88 | 5.70 | 5.51 |
| **RVPMDL** | 544 | 457 | 483 | 463 |
| **RVPA** | .86 | .90 | .91 | .94 |
| **RVPPH** | .47 | .63 | .66 | .74 |
| **RVPPFA** | .007 | .007 | .007 | .003 |
| **RVPTM** | 28.7 | 20.0 | 18.30 | 14.00 |
| **SSTSSRT** | 276 | 251 | 231 | 216 |
| **SSTDEG** | 2.42 | 1.91 | .90 | 1.12 |
| **SSTDES** | 41.70 | 41.20 | 36.20 | 35.80 |
| **SSTMT** | 7.50 | 4.47 | 15.90 | 4.41 |
| **CADSS** | 11.00 | 6.20 | 4.09 | 4.04 |
| **Positive Affect** | 23.60 | 33.90 | 25.30 | 28.20 |
| **Negative Affect** | 16.10 | 11.80 | 14.60 | 12.40 |
| **Baseline Heartrate** | 74.60 | 69.20 | 73.80 | 65.90 |
| **Baseline EDA** | 10.19 | 9.98 | 12.81 | 10.07 |
| **Baseline RMSSD** | 65.00 | 72.80 | 37.00 | 59.20 |
| **Arousal** |  |  |  |  |
| *NeutralWatch* | 4.25 | 4.13 | 3.54 | 3.04 |
| *NegativeWatch* | 5.62 | 5.14 | 5.30 | 4.26 |
| *NegativeDampen* | 5.66 | 5.17 | 5.22 | 4.32 |
| **Valence** |  |  |  |  |
| *NeutralWatch* | 5.11 | 5.37 | 4.99 | 5.42 |
| *NegativeWatch* | 3.71 | 4.29 | 3.77 | 4.29 |
| *NegativeDampen* | 3.68 | 4.38 | 3.48 | 4.24 |
| **Heartrate** |  |  |  |  |
| *NeutralWatch* | 70.30 | 68.70 | 70.30 | 65.80 |
| *NegativeWatch* | 69.50 | 68.00 | 68.80 | 65.40 |
| *NegativeDampen* | 69.60 | 68.50 | 68.60 | 65.80 |
| **EDA** |  |  |  |  |
| *NeutralWatch* | 11.02 | 8.90 | 15.83 | 11.07 |
| *NegativeWatch* | 11.12 | 8.74 | 16.00 | 11.15 |
| *NegativeDampen* | 10.90 | 9.08 | 15.58 | 11.10 |
| **RMSSD** |  |  |  |  |
| *NeutralWatch* | 42.80 | 54.70 | 80.20 | 29.40 |
| *NegativeWatch* | 45.10 | 49.90 | 43.20 | 26.60 |
| *NegativeDampen* | 61.30 | 104.20 | 29.00 | 46.60 |

*Notes.* HTT = heartbeat tracking task; RVP = rapid visual information processing task; RVPMDL = RVP median response latency; RVPA = RVP sensitivity to target sequence; RVPPH = RVP probability of hit; RVPPFA = RVP probability of false alarm; RVPTM = RVP total misses; SST = stop signal task; SSTSSRT = stop signal reaction time; SSTDEG = SST direction errors go trials; SSTDES = SST direction errors stop trials; SSTMT = SST missed trials; CADSS = clinician administered dissociative states scale; EDA = electrodermal activity; RMSSD = root mean square of successive differences.

**Table S8.** Linear mixed effects models pre-post six weeks.

|  | ***ß*** | **SE** | **95% CI** | **t** | **p** |
| --- | --- | --- | --- | --- | --- |
| **HTT Accuracy** |  |  |  |  |  |
| *Intercept* | .36 | .27 | -.16, .87 | 1.31 | .20 |
| *Group* | -.04 | .17 | -.38, .29 | -.25 | .81 |
| *Time* | .11 | .10 | -.09, .30 | 1.06 | .31 |
| *Group x Time* | -.03 | .07 | -.16, .10 | -.42 | .68 |
| **HTT Confidence** |  |  |  |  |  |
| *Intercept* | 1.26 | 2.41 | -3.36, 5.87 | .52 | .60 |
| *Group* | 2.34 | 1.55 | -.62, 5.32 | 1.51 | .14 |
| *Time* | .82 | .72 | -.62, 2.22 | 1.14 | .27 |
| *Group x Time* | -.53 | .48 | -1.45, .44 | -1.10 | .28 |
| **RVPMDL** |  |  |  |  |  |
| *Intercept* | 692.49 | 119.00 | 464.19, 920.12 | 5.82 | .00 |
| *Group* | -108.28 | 74.62 | -251.28, 34.72 | -1.45 | .16 |
| *Time* | -88.04 | 52.03 | -188.39, 13.61 | -1.69 | .11 |
| *Group x Time* | 47.60 | 33.54 | -17.49, 112.64 | 1.42 | .17 |
| **RVPA** |  |  |  |  |  |
| *Intercept* | .77 | .04 | .70, .84 | 21.59 | .00 |
| *Group* | .07 | .02 | .03, .11 | 3.02 | .00 |
| *Time* | .05 | .01 | .03, .07 | 4.83 | .00 |
| *Group x Time* | -.02 | .01 | -.03, -.01 | -3.15 | .01 |
| **RVPPH** |  |  |  |  |  |
| *Intercept* | .08 | .14 | -.19, .34 | .57 | .57 |
| *Group* | .28 | .09 | .11, .44 | 3.20 | .00 |
| *Time* | .20 | .04 | .13, .27 | 5.35 | .00 |
| *Group x Time* | -.09 | .02 | -.13, -.04 | -3.57 | .00 |
| **RVPPFA** |  |  |  |  |  |
| *Intercept* | .00 | .01 | -.01, .02 | .36 | .73 |
| *Group* | .00 | .00 | -.01, .01 | .57 | .58 |
| *Time* | .00 | .00 | -.00, .01 | .92 | .37 |
| *Group x Time* | -.00 | .00 | -.01, .00 | -.99 | .33 |
| **RVPTM** |  |  |  |  |  |
| *Intercept* | 49.77 | 7.40 | 35.67, 64.01 | 6.73 | .00 |
| *Group* | -14.98 | 4.68 | -23.99, -6.07 | -3.20 | .00 |
| *Time* | -10.89 | 2.04 | -14.81, -6.84 | -5.35 | .00 |
| *Group x Time* | 4.68 | 1.31 | 2.08, 7.21 | 3.57 | .00 |
| **SSTSSRT** |  |  |  |  |  |
| *Intercept* | 331.98 | 41.41 | 252.95, 410.91 | 8.02 | .00 |
| *Group* | -48.29 | 27.08 | -99.91, 3.37 | -1.78 | .08 |
| *Time* | -11.01 | 16.13 | -42.11, 20.86 | -.68 | .50 |
| *Group x Time* | 3.45 | 10.78 | -17.68, 24.28 | .32 | .75 |
| **SSTDEG** |  |  |  |  |  |
| *Intercept* | 5.57 | 2.12 | 1.48, 9.63 | 2.63 | .01 |
| *Group* | -2.45 | 1.39 | -5.09, .23 | -1.76 | .09 |
| *Time* | -1.62 | .94 | -3.44, .23 | -1.72 | .10 |
| *Group x Time* | .91 | .62 | -.32, 2.11 | 1.46 | .16 |
| **SSTDES** |  |  |  |  |  |
| *Intercept* | 48.20 | 2.85 | 42.77, 53.69 | 16.93 | .00 |
| *Group* | -6.53 | 1.86 | -10.14, -2.98 | -3.51 | .00 |
| *Time* | -1.05 | 1.17 | -3.49, 1.18 | -.90 | .38 |
| *Group x Time* | 1.05 | .78 | -.43, 2.70 | 1.35 | .20 |
| **SSTMT** |  |  |  |  |  |
| *Intercept* | 4.11 | 11.20 | -17.38, 25.85 | .37 | .72 |
| *Group* | 5.86 | 7.25 | -8.24, 19.72 | .81 | .43 |
| *Time* | -4.03 | 2.64 | -9.08, 1.38 | -1.53 | .15 |
| *Group x Time* | 1.56 | 1.78 | -2.18, 4.96 | .87 | .40 |
| **CADSS** |  |  |  |  |  |
| *Intercept* | 21.11 | 5.68 | 10.26, 31.94 | 3.71 | .00 |
| *Group* | -8.00 | 3.67 | -14.99, -.99 | -2.18 | .04 |
| *Time* | -3.20 | 2.29 | -7.63, 1.30 | -1.40 | .18 |
| *Group x Time* | 1.09 | 1.54 | -1.93, 4.05 | .71 | .49 |
| **Positive Affect** |  |  |  |  |  |
| *Intercept* | 13.71 | 7.06 | .24, 27.17 | 1.94 | .06 |
| *Group* | 5.06 | 4.56 | -3.63, 13.76 | 1.11 | .27 |
| *Time* | 8.19 | 2.58 | 3.05, 13.20 | 3.17 | .01 |
| *Group x Time* | -3.37 | 1.74 | -6.71, .17 | -1.94 | .07 |
| **Negative Affect** |  |  |  |  |  |
| *Intercept* | 17.65 | 4.67 | 8.71, 26.63 | 3.78 | .00 |
| *Group* | -1.21 | 3.02 | -7.02, 4.56 | -.40 | .69 |
| *Time* | -.12 | 2.10 | -4.24, 3.93 | -.06 | .95 |
| *Group x Time* | -.23 | 1.40 | -2.92, 2.51 | -.17 | .87 |
| **Baseline Heartrate** |  |  |  |  |  |
| *Intercept* | 77.68 | 8.74 | 60.99, 94.36 | 8.89 | .00 |
| *Group* | -2.01 | 5.63 | -12.74, 8.73 | -.36 | .72 |
| *Time* | -2.34 | 3.09 | -8.39, 3.67 | -.76 | .46 |
| *Group x Time* | 1.22 | 2.04 | -2.77, 5.20 | .60 | .56 |
| **Baseline EDA** |  |  |  |  |  |
| *Intercept* | 8.08 | 5.57 | -2.53, 18.69 | 1.45 | .16 |
| *Group* | 3.33 | 3.69 | -3.70, 10.37 | .90 | .37 |
| *Time* | -.79 | 2.13 | -4.93, 3.36 | -.37 | .72 |
| *Group x Time* | -.44 | 1.43 | -3.22, 2.34 | -.31 | .76 |
| **Baseline RMSSD** |  |  |  |  |  |
| *Intercept* | 84.71 | 51.64 | -13.93, 183.22 | 1.64 | .11 |
| *Group* | -25.30 | 33.25 | -88.72, 38.21 | -.76 | .45 |
| *Time* | 8.21 | 20.42 | -31.21, 48.35 | .40 | .69 |
| *Group x Time* | -2.66 | 13.46 | -29.13, 23.30 | -.20 | .85 |
| **Arousal** |  |  |  |  |  |
| *Intercept* | 3.55 | .33 | 2.92, 4.18 | 10.77 | .00 |
| *Group: Yoga* | .67 | .46 | -.21, 1.55 | 1.47 | .15 |
| *Time: Post* | .19 | .40 | -.57, .97 | .49 | .63 |
| *Condition: NegativeWatch* | 1.74 | .37 | 1.02, 2.45 | 4.70 | .00 |
| *Condition: NegativeDampen* | 1.64 | .37 | .92, 2.36 | 4.42 | .00 |
| *Group x Time: Yoga x Post* | -.61 | .53 | -1.66, .42 | -1.15 | .25 |
| *GroupxCondition: Yoga x NegativeWatch* | -.32 | .51 | -1.31, .67 | -.64 | .53 |
| *Group x Condition: Yoga x NegativeDampen* | -.20 | .51 | -1.19, .78 | -.40 | .69 |
| *Time x Condition: Post x NegativeWatch* | -.52 | .57 | -1.62, .58 | -.91 | .36 |
| *Time x Condition: Post x NegativeDampen* | -.39 | .56 | -1.48, .70 | -.69 | .49 |
| *Group x Time x Condition: Yoga x Post x NegativeWatch* | .53 | .76 | -.95, 2.01 | .70 | .48 |
| *Group x Time x Condition: Yoga x Post x NegativeDampen* | -.21 | .75 | -1.67, 1.26 | -.28 | .78 |
| **Valence** |  |  |  |  |  |
| *Intercept* | 5.03 | .26 | 4.53, 5.53 | 19.53 | .00 |
| *Group: Yoga* | .06 | .36 | -.63, .76 | .18 | .86 |
| *Time: Post* | .51 | .33 | -.14, 1.15 | 1.54 | .12 |
| *Condition: NegativeWatch* | -1.27 | .30 | -1.86, -.69 | -4.22 | .00 |
| *Condition: NegativeDampen* | -1.54 | .30 | -2.12, -.95 | -5.12 | .00 |
| *Group x Time: Yoga x Post* | -.13 | .45 | -1.01, .74 | -.30 | .77 |
| *GroupxCondition: Yoga x NegativeWatch* | -.11 | .42 | -.94, ,71 | -.26 | .79 |
| *Group x Condition: Yoga x NegativeDampen* | .13 | .42 | -.68, .95 | -.32 | .75 |
| *Time x Condition: Post x NegativeWatch* | -.04 | .47 | -.95, .87 | -.08 | .93 |
| *Time x Condition: Post x NegativeDampen* | .17 | .46 | -.72, 1.06 | -.37 | .72 |
| *Group x Time x Condition: Yoga x Post x NegativeWatch* | -.47 | .64 | -1.71, .77 | -.74 | .46 |
| *Group x Time x Condition: Yoga x Post x NegativeDampen* | -.02 | .63 | -1.20, 1.24 | -.03 | .98 |
| **Heartrate** |  |  |  |  |  |
| *Intercept* | 68.60 | 2.69 | 63.35, 73.86 | 25.47 | .00 |
| *Group: Yoga* | 1.72 | 3.73 | -5.55, 9.00 | .46 | .65 |
| *Time: Post* | .46 | .96 | -1.40, 2.32 | .48 | .63 |
| *Condition: NegativeWatch* | .20 | .85 | -1.44, 1.85 | .24 | .81 |
| *Condition: NegativeDampen* | -.01 | .85 | -1.66, 1.64 | -.01 | .99 |
| *Group x Time: Yoga x Post* | -2.26 | 1.30 | -4.77, .26 | -1.74 | .08 |
| *GroupxCondition: Yoga x NegativeWatch* | -1.09 | 1.18 | -3.38, 1.20 | -.93 | .35 |
| *Group x Condition: Yoga x NegativeDampen* | -.81 | 1.18 | -3.10, 1.48 | -.69 | .49 |
| *Time x Condition: Post x NegativeWatch* | -1.17 | 1.33 | -3.75, 1.41 | -.88 | .38 |
| *Time x Condition: Post x NegativeDampen* | -.80 | 1.32 | -3.37, 1.77 | -.61 | .54 |
| *Group x Time x Condition: Yoga x Post x NegativeWatch* | 1.76 | 1.80 | -1.74, 5.26 | .98 | .33 |
| *Group x Time x Condition: Yoga x Post x NegativeDampen* | .12 | 1.80 | -3.37, 3.61 | .07 | .95 |
| **EDA** |  |  |  |  |  |
| *Intercept* | 15.43 | 2.14 | 11.26, 19.61 | 7.21 | .00 |
| *Group: Yoga* | -4.42 | 2.96 | -10.19, 1.35 | -1.49 | .15 |
| *Time: Post* | -5.04 | .70 | -6.39, -3.68 | -7.21 | .00 |
| *Condition: NegativeWatch* | .17 | .63 | -1.05, 1.39 | .27 | .79 |
| *Condition: NegativeDampen* | -.36 | .63 | -1.59, .87 | -.57 | .57 |
| *Group x Time: Yoga x Post* | 2.58 | .93 | .77, 4.38 | 2.77 | .01 |
| *GroupxCondition: Yoga x NegativeWatch* | -.08 | .86 | -1.74, 1.58 | -.09 | .92 |
| *Group x Condition: Yoga x NegativeDampen* | .23 | .86 | -1.44, 1.90 | .27 | .79 |
| *Time x Condition: Post x NegativeWatch* | -.27 | .95 | -2.11, 1.58 | -.28 | .78 |
| *Time x Condition: Post x NegativeDampen* | .11 | .95 | -1.74, 1.95 | .11 | .91 |
| *Group x Time x Condition: Yoga x Post x NegativeWatch* | .08 | 1.28 | -2.41, 2.56 | .06 | .95 |
| *Group x Time x Condition: Yoga x Post x NegativeDampen* | -.20 | 1.28 | -2.68, 2.28 | -.16 | .88 |
| **RMSSD** |  |  |  |  |  |
| *Intercept* | 50.90 | 15.03 | 21.98, 79.85 | 3.39 | .00 |
| *Group: Yoga* | 1.01 | 20.56 | -38.59, 40.57 | .05 | .96 |
| *Time: Post* | 17.10 | 15.98 | -13.95, 48.15 | 1.07 | .29 |
| *Condition: NegativeWatch* | 22.93 | 15.99 | -8.19, 53.95 | 1.43 | .15 |
| *Condition: NegativeDampen* | -22.02 | 16.44 | -53.82, 10.10 | -1.34 | .18 |
| *Group x Time: Yoga x Post* | -6.42 | 21.94 | -49.04, 36.19 | -.29 | .77 |
| *GroupxCondition: Yoga x NegativeWatch* | -12.38 | 22.00 | -55.09, 30.41 | -.56 | .57 |
| *Group x Condition: Yoga x NegativeDampen* | 8.69 | 22.11 | -34.50, 51.46 | .39 | .69 |
| *Time x Condition: Post x NegativeWatch* | -57.54 | 23.04 | -102.11, -12.54 | -2.50 | .01 |
| *Time x Condition: Post x NegativeDampen* | -10.37 | 23.17 | -55.39, 34.66 | -.45 | .65 |
| *Group x Time x Condition: Yoga x Post x NegativeWatch* | 22.92 | 31.31 | -38.19, 83.52 | .73 | .46 |
| *Group x Time x Condition: Yoga x Post x NegativeDampen* | -6.30 | 31.14 | -66.80, 54.18 | -.20 | .84 |

*Notes.* HTT = heartbeat tracking task; RVP = rapid visual information processing task; RVPMDL = RVP median response latency; RVPA = RVP sensitivity to target sequence; RVPPH = RVP probability of hit; RVPPFA = RVP probability of false alarm; RVPTM = RVP total misses; SST = stop signal task; SSTSSRT = stop signal reaction time; SSTDEG = SST direction errors go trials; SSTDES = SST direction errors stop trials; SSTMT = SST missed trials; CADSS = clinician administered dissociative states scale; EDA = electrodermal activity; RMSSD = root mean square of successive differences.

**Table S9.** Three-way mixed ANOVAs pre-post six-week programme.

| **Variable** | **ANOVA** | **F (df)** | **p** | ***ηp2*** |
| --- | --- | --- | --- | --- |
| Arousal | Group x Time x Condition | .49 (2, 428.74) | .61 | .00 |
| Valence | Group x Time x Condition | .38 (2, 431.63) | .69 | .00 |
| Heart rate | Group x Time x Condition | .62 (2, 452.03) | .54 | .00 |
| Skin conductance | Group x Time x Condition | .03 (2, 430.95) | .98 | .00 |
| RMSSD | Group x Time x Condition | 3.02 (2, 452.28) | .05 | .01 |

*Notes.* RMSSD = root mean square of successive differences.

**Table S10.** Laboratory measures by group across time post six-week programme (emmeans).

|  | **Yoga** |  | **Music** |  |
| --- | --- | --- | --- | --- |
|  | **Baseline** | **Post 6 weeks** | **Baseline** | **Post 6 weeks** |
| **HTT Accuracy** | .39 | .55 | .32 | .42 |
| **HTT Confidence** | 3.90 | 4.48 | 5.70 | 5.23 |
| **RVPMDL** | 544 | 463 | 483 | 497 |
| **RVPA** | .86 | .92 | .91 | .93 |
| **RVPPH** | .47 | .70 | .66 | .72 |
| **RVPPFA** | .007 | .008 | .007 | .005 |
| **RVPTM** | 28.60 | 16.20 | 18.30 | 15.20 |
| **SSTSSRT** | 276 | 261 | 231 | 223 |
| **SSTDEG** | 2.42 | .99 | .88 | 1.28 |
| **SSTDES** | 41.70 | 41.70 | 36.20 | 38.30 |
| **SSTMT** | 7.50 | 2.57 | 14.91 | 13.10 |
| **CADSS** | 11.00 | 6.77 | 4.09 | 2.04 |
| **Positive Affect** | 23.60 | 33.20 | 25.30 | 28.20 |
| **Negative Affect** | 16.10 | 15.40 | 14.60 | 13.50 |
| **Baseline Heartrate** | 74.60 | 72.30 | 73.80 | 74.00 |
| **Baseline EDA** | 10.19 | 7.75 | 13.09 | 9.77 |
| **Baseline RMSSD** | 65.00 | 76.10 | 37.00 | 42.80 |
| **Arousal** |  |  |  |  |
| *NeutralWatch* | 4.22 | 3.80 | 3.55 | 3.74 |
| *NegativeWatch* | 5.64 | 5.23 | 5.29 | 4.96 |
| *NegativeDampen* | 5.66 | 4.64 | 5.19 | 4.99 |
| **Valence** |  |  |  |  |
| *NeutralWatch* | 5.09 | 5.47 | 5.03 | 5.54 |
| *NegativeWatch* | 3.71 | 3.58 | 3.76 | 4.23 |
| *NegativeDampen* | 3.69 | 4.25 | 3.49 | 4.17 |
| **Heartrate** |  |  |  |  |
| *NeutralWatch* | 70.30 | 68.50 | 68.60 | 69.10 |
| *NegativeWatch* | 69.40 | 68.20 | 68.80 | 68.10 |
| *NegativeDampen* | 69.50 | 67.00 | 68.60 | 68.30 |
| **EDA** |  |  |  |  |
| *NeutralWatch* | 11.01 | 8.56 | 15.43 | 10.40 |
| *NegativeWatch* | 11.10 | 8.45 | 15.60 | 10.30 |
| *NegativeDampen* | 10.88 | 8.33 | 15.07 | 10.14 |
| **RMSSD** |  |  |  |  |
| *NeutralWatch* | 51.90 | 62.60 | 50.90 | 68.00 |
| *NegativeWatch* | 62.50 | 38.50 | 73.80 | 33.40 |
| *NegativeDampen* | 38.60 | 32.60 | 28.90 | 35.60 |

*Notes.* HTT = heartbeat tracking task; RVP = rapid visual information processing task; RVPMDL = RVP median response latency; RVPA = RVP sensitivity to target sequence; RVPPH = RVP probability of hit; RVPPFA = RVP probability of false alarm; RVPTM = RVP total misses; SST = stop signal task; SSTSSRT = stop signal reaction time; SSTDEG = SST direction errors go trials; SSTDES = SST direction errors stop trials; SSTMT = SST missed trials; CADSS = clinician administered dissociative states scale; EDA = electrodermal activity; RMSSD = root mean square of successive differences.
